# Somatic mutations reveal hyperactive Notch signaling and racial disparities in prurigo nodularis

**DOI:** 10.1101/2023.09.25.23295810

**Authors:** Ahmad Rajeh, Hannah L. Cornman, Anuj Gupta, Mindy D. Szeto, Anusha Kambala, Olusola Oladipo, Varsha Parthasarathy, Junwen Deng, Sarah Wheelan, Thomas Pritchard, Madan M. Kwatra, Yevgeniy R. Semenov, Alexander Gusev, Srinivasan Yegnasubramanian, Shawn G. Kwatra

## Abstract

Prurigo nodularis (PN) is a chronic inflammatory skin disease that disproportionately affects African Americans and is characterized by pruritic skin nodules of unknown etiology. Little is known about genetic alterations in PN pathogenesis, especially relating to somatic events which are often implicated in inflammatory conditions. We thus performed whole-exome sequencing on 54 lesional and nonlesional skin biopsies from 17 PN patients and 10 atopic dermatitis (AD) patients for comparison. Somatic mutational analysis revealed that PN lesional skin harbors pervasive somatic mutations in fibrotic, neurotropic, and cancer-associated genes. Nonsynonymous mutations were most frequent in *NOTCH1* and the Notch signaling pathway, a regulator of cellular proliferation and tissue fibrosis, and *NOTCH1* mutations were absent in AD. Somatic copy-number analysis, combined with expression data, showed that recurrently deleted and downregulated genes in PN lesional skin are associated with axonal guidance and extension. Follow-up immunofluorescence validation demonstrated increased *NOTCH1* expression in PN lesional skin fibroblasts and increased Notch signaling in PN lesional dermis. Finally, multi-center data revealed a significantly increased risk of *NOTCH1-* associated diseases in PN patients. In characterizing the somatic landscape of PN, we uncover novel insights into its pathophysiology and identify a role for dysregulated Notch signaling in PN.

## Introduction

Prurigo nodularis (PN) is a chronic inflammatory skin disease characterized by intensely pruritic, hyperkeratotic skin nodules on the trunk and extremities (1, 2). Compared to more common and better characterized chronic pruritic dermatoses like atopic dermatitis (AD) or psoriasis, PN is associated with greater itch intensity (3), as well as a significant quality of life impairment (4, 5). PN emerges in middle age, disproportionately affects African Americans, and is associated with multiple systemic conditions (6–9). Despite this significant clinical burden, the etiology of PN remains poorly understood.

The current understanding of PN biology centers around an interplay between cutaneous inflammation, neuronal dysregulation, and altered keratinocyte differentiation and fibroblast signaling (10–13). Recent transcriptomic studies show characteristic patterns of immune polarization in PN patients, including both Th2/Th17-centered cutaneous immune activation, and cutaneous and systemic Th22-related cytokine upregulation (10, 14). Black patients have a greater genetic risk of developing PN and distinct inflammatory signatures are seen in African American PN patients, suggesting the existence of multiple disease endotypes (15–17). However, to which extent those patterns might be explained by genetic variation or environmental exposures remains unknown. In better studied chronic pruritic dermatoses such as AD and psoriasis, genomic studies have accelerated our understanding of disease pathology and informed new treatments (18, 19), but similar investigations are lacking for PN.

The relevance of genomic studies in inflammatory skin disease includes postzygotic variation. Somatic mutations throughout the body are known to drive neoplasms, but growing evidence also points to clonal expansions harboring somatic mutations in non-neoplastic disease and healthy-appearing tissue, including the skin (20–23). Such findings have informed our understanding of both the disease and the corresponding tissue biology. For example, colonic mucosa of patients with inflammatory bowel disease displays positive selection for mutations in interleukin-17 pathway genes, which may confer a protective advantage to mucosal epithelia (20, 24). Thus, delineating somatic events associated with PN may further our understanding of not only disease pathology, but also cutaneous molecular adaptations in the setting of chronic itch, fibrosis, and neuroinflammation. This is especially pressing given the comorbidities in PN patients, including a higher risk of skin and internal malignancies (25, 26), which remain largely unexplained.

In this study, we characterize the landscape of somatic events in PN lesional skin. We perform whole-exome sequencing (WES) on individual-matched lesional and nonlesional skin biopsies from a diverse cohort of PN patients, as well as AD patients as a reference group. We explore the mutational landscape of PN at the individual nodule level, identifying somatic events in lesional PN skin compared to adjacent healthy-appearing skin. We also contrast the somatic profile of PN to that of AD to elucidate molecular pathways specific to PN. Our somatic analysis also leads us to functional and multi-center epidemiological follow-up investigations. To our knowledge, this work represents the first and largest genomic study of PN.

## Results

### Whole-exome analysis

An illustration of the study design is shown (Figure 1A). Briefly, we recruited 17 patients from the Johns Hopkins Itch Center diagnosed with PN (Figure 1B and C) fulfilling our selection criteria (see Methods). Patient demographics are summarized in Figure 1D. Two skin punch-biopsies were obtained from each patient, one from a prurigo nodule (lesional) and one from normal-appearing skin (nonlesional) within 10 cm of the nodule. Since high-throughput WES can provide sufficient resolution for the detection of somatic events, we generated WES data from 17 lesional and 17 nonlesional PN samples in this study. In the same method, we also generated 20 matched WES datasets from 10 patients with AD for use as reference group (Figure 1A and D).

**Figure 1.**
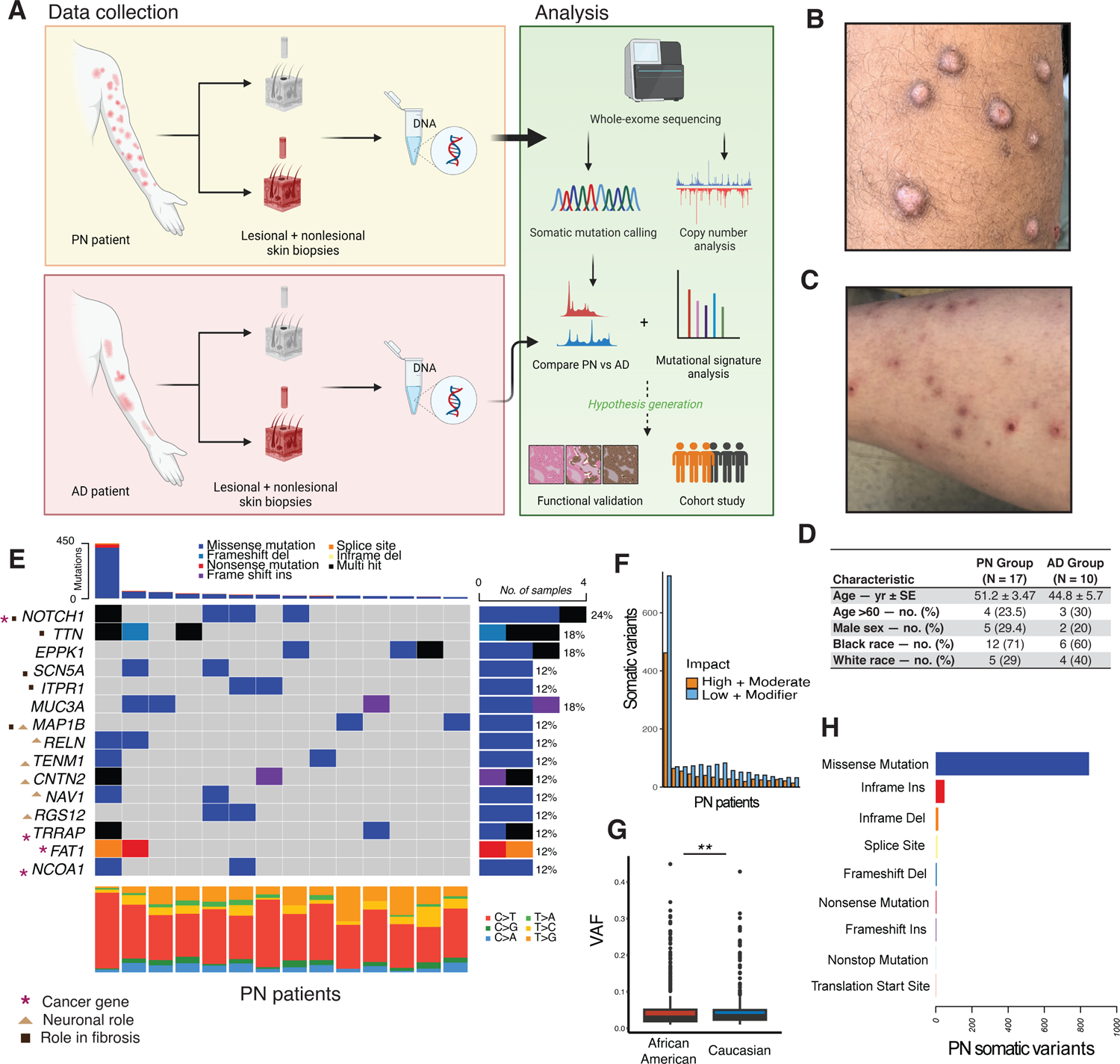
Somatic mutational landscape of PN. (A) Schematic of the study design including sample collection and analysis. (B, C) Example skin images of two patients with prurigo nodularis (PN) enrolled in this study. (B) Dorsal left arm of an African American patient with scattered prurigo nodules. (C) Right arm of a Caucasian patient with scattered prurigo nodules. (D) Demographic information of PN and atopic dermatitis (AD) patients. (E) Waterfall plot displaying somatic mutations in 15 of the most frequently mutated genes in PN lesional skin compared to nonlesional skin. Ties were broken by known gene function. Columns represent the 17 PN patients. Numbers shown on the right indicate the frequency of gene mutation. Total nonsynonymous somatic SNPs and indels per sample are displayed on top. One sample was hypermutated with 450 mutations. Bottom plot shows the frequency of different classes of SNVs. Variants annotated as Multi Hit are those genes which mutated more than once in the same sample. (F) Total number of somatic mutations per sample, broken down by predicted impact on protein product based on snpEff, with functional (High or Moderate) impact in yellow and nonfunctional (Low or Modifier) in blue. (G) Boxplot showing the variant allele frequency (VAF) of nonsynonymous somatic mutations across race. (H) **, *P* < 0.01.

From our PN cohort (34 WES datasets across 17 patients), we obtained approximately 1.7 billion reads (mean and range = 4.9 × 10^7^ [4.2 × 10^7^ to 6.1 × 10^7^] and 5.2 × 10^7^ [4.3 × 10^7^ to 7.0 × 10^7^] for lesional and nonlesional samples, respectively). After quality control and alignment to the hg38 human reference genome, we noted an average sequencing coverage of 195 (187 and 202 for lesional and nonlesional, respectively). On average, more than 93% of the exome was sequenced to at least 20X depth, providing a reliable signal for variant calling (27). Detailed sequencing and alignment information, including results from preprocessing and variant calling on our AD control cohort, is provided (Table S1). One pair of AD lesional/nonlesional samples was excluded from downstream analyses because the nonlesional sample had less than 50% of the exome sequenced to at least 20X depth.

### Somatic variation in PN lesional skin

Somatic variants in PN nodules were identified by matching PN lesional to nonlesional samples per patient. The landscape of somatic mutations in PN is shown in Figure 1E-H. Following quality control, we identified 2387 high-confidence somatic single nucleotide variations (SNVs) SNVs and small insertions/deletions (indels) in PN lesional skin affecting 1933 genes. Mutational analysis showed a median of 75 somatic (lesion-specific) mutations per patient, including SNVs and indels. We observed a median mutational burden of approximately 0.52 per megabase of coding region (range of 0.26 to 9 per megabase; outlier sample was further characterized in Figure S1). We noted 35% (847 of 2387), 2.0% (48 of 2387), 0.59% (14 of 2387), 0.58% (13 of 2387), and 0.3% (8 of 2387) missense, nonsense, splice site altering, in-frame indels, and frameshift indels, respectively. After predicting the effects of variants on protein function, variants were classified into significance categories “High + Moderate”, indicating likely change in protein sequence, and “Low + Modifier”, indicating no known change in protein sequence (Figure 1F). We found a median of 29 functional, lesion-specific somatic SNVs or indels per PN patient (range of 13 to 450) (Figure 1F).

We observed a median somatic variant allele frequency (VAF) of 0.031 (range of 0.01 to 0.449) in PN lesional skin, with Caucasian PN patients displaying a higher VAF than African American PN patients (0.033 compared to 0.029, *P* = 0.0056, *Wilcoxon*) (Figure 1G and H). On assessment of somatic variants at the gene level, there were 46 genes with nonsynonymous somatic mutations in at least two out of 17 patients with PN. We grouped recurrently-mutated genes using three *a priori*-defined gene sets relevant to PN based on literature and clinical judgement. There were 5 genes associated with pathologic fibrosis (*NOTCH1*, *SCN5A*, *MAP1B, TTN*, *ITPR1*) (28), 6 genes associated with neuronal migration or projection (*MAP1B*, *RELN*, *TENM1*, *CNTN2, NAV1, RGS12*) by gene ontology, and 4 cancer-associated genes (*NOTCH1*, *TRRAP, FAT1*, *NCOA1*) according to the Cancer Gene Census (CGC) from the Catalogue of Somatic Mutations in Cancer (COSMIC) (29). *NOTCH1* had the most frequent nonsynonymous somatic mutations: we found 4 missense SNVs (p.Arg1962His, p.Asn70Ile, p.Glu450Lys, and p.Asn325Lys) and 1 in-frame deletion (p.Val413-Asp414del) across 4 patients, 2 of whom were African American. *NOTCH1* is an intracellular regulator of the Notch family with pleiotropic functions in cellular proliferation and tissue fibrosis, and a known hallmark driver of cutaneous squamous cell carcinoma (cSCC) (30, 31). 3 out of 5 identified *NOTCH1* mutations where within the extracellular epidermal growth factor (EGF)-like domain, which is involved in ligand binding and preventing constitutional activation (32). Somatic mutations in *NOTCH1* had a significant co-occurrence with mutations in *NCOA1, MISP*, *NAV1*, *MYO1C*, *RGS12*, and *VPS13B* (*P* < 0.05, pairwise *Fisher’s exact test*) (Figure 2A).

**Figure 2.**
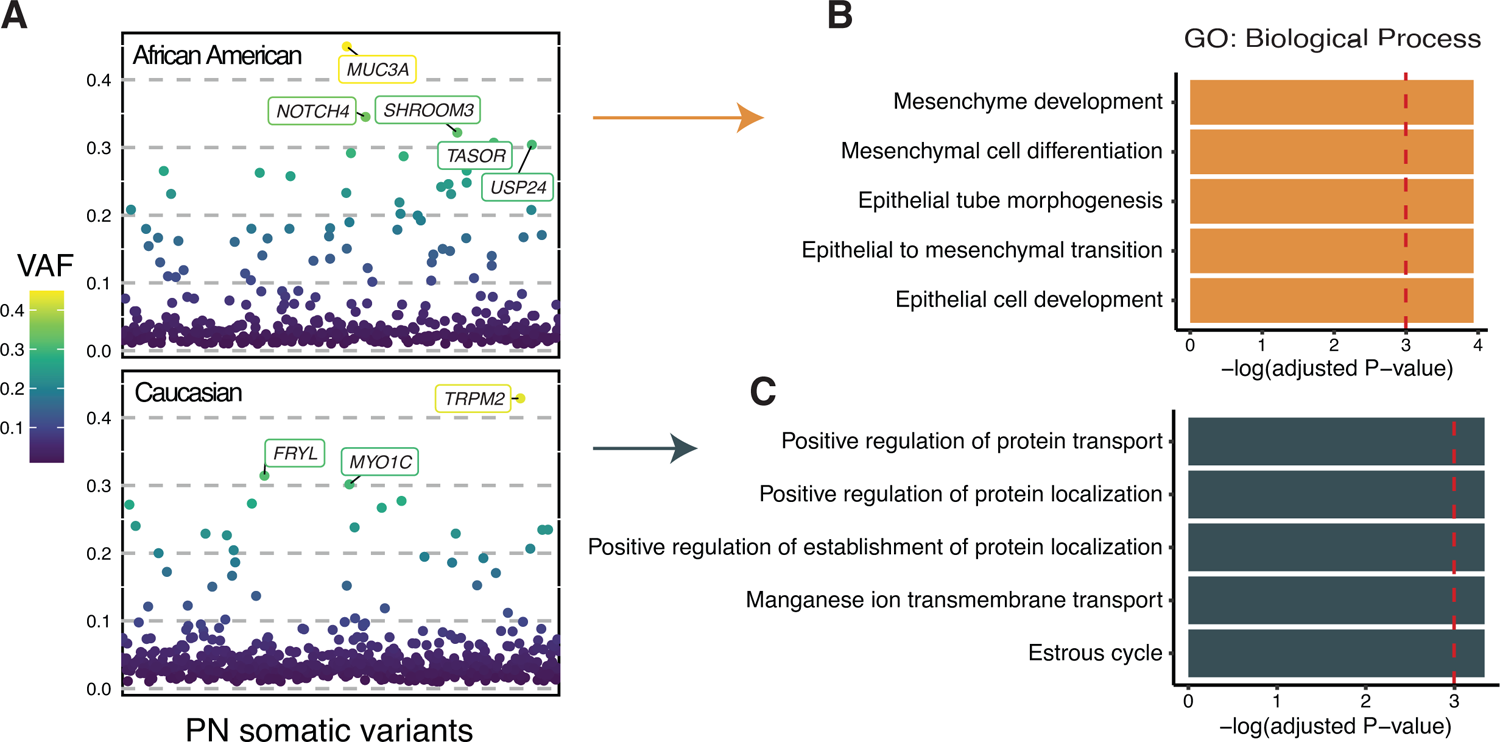
Distinct somatic selection in lesional skin of African American patients with PN. (A) Nonsynonymous somatic mutations are ordered by their genomic locations on the x-axis and the corresponding VAF is shown on the y-axis. Variants with 0.3 or higher VAF are labelled with their gene name. (B and C) Bar graph showing the pathway enrichment results of high-VAF (>0.3) nonsynonymous mutations in African American and Caucasian PN patients, respectively.

#### Genes associated with fibrosis

In addition to *NOTCH1*, we identified somatic mutations in other profibrotic genes. We noted two missense mutations in *SCN5A* (p.Gly1158Asp, p.Arg1638Gln) across two PN patients. *SCN5A* encodes the alpha subunit of the Na_v_1.5 sodium channel, which plays a key role in the depolarization of cardiomyocytes and whose channelopathy is linked to cardiac fibrosis (33). *SCN10A* from the same gene family, which encodes Na_v_1.8, which regulates pro-inflammatory responses in the skin and was significantly upregulated in the epidermis of rosacea and psoriasis skin lesions (34).

We found two missense mutations in *ITPR1* (p.Ala805Val, p.Glu914Lys) affecting two PN patients. *ITPR1* encodes one of the three members of the IP_3_-receptor family that form calcium release channels and is associated with pancreatic fibrosis (28), in addition to chronic itch mediated by astrocytes (35). We also observed two recurrent missense mutations in *MAP1B* (p.Val1549Gly x 2) across two patients. While *MAP1B* is thought to be primarily involved in microtubule assembly and neurogenesis, it also plays a role in fibrosis, particularly in the eye (28, 36).

In addition, we found *TTN* mutated in 3 patients including 4 missense mutations (p.Ala25524Val, p.Asn8023Lys, p.Ala35193Thr, and p.Val7022Ala) and one frame-shift deletion (p.Val29546AsnfsTer3). *TTN* has an established role in interstitial fibrosis and is frequently mutated in numerous cancers, but it encodes the largest protein in humans and is frequently mutated in reference populations (37, 38). Of note, we found a recurrent p.Ile1690Val missense mutation in Epiplakin 1 (*EPPK1*) in 3 of the PN patients. While *EPPK1* was not in our pre-defined gene sets, this was the only gene with 3 or more identical recurrent somatic mutations, at either the DNA or amino acid level. *EPPK1* encodes a member of the plakin family which is involved in the organization of cytoskeletal architecture and has been shown to accelerate keratinocyte migration during wound healing (39, 40). We also found several mutations in genes related to neuronal migration or projection, detailed below.

#### Neurotropic genes

The mutations in *MAP1B* are described above since it is found to have both fibrotic as well as neurotropic functions (28, 36). *RELN*, which is involved in nerve migration, projection, and synaptic plasticity was found to have two missense mutations (p.Thr468Pro, p.Val167Leu) across two patients. We also found two missense mutations across two patients in *TENM1* (p.Ala2661Ser, p.Gly1134Arg) and *CNTN2* (p.Ala2661Ser, p.Gly1134Arg). *TENM1* encodes a protein of the teneurin subfamily which is thought to function as a neuronal cellular signal transducer and is one of the most highly mutated genes in melanoma (41). *CNTN2* encodes contactin 2, which is found to play an important role in neuronal excitability (42). In addition, we noted two missense mutations across two patients in *RGS12* (p.Gly454Asp, p.Arg403Cys) and *NAV1* (p.Pro1507Ser, p.Arg1581Cys)*. RGS12* was shown to be a critical modulator of serotonergic neurotransmission (43), while *NAV1* is a relatively understudied gene that is involved in neuronal development and regeneration (44).

#### Cancer-associated genes

To identify mutations in known cancer genes, we overlapped our set of 46 genes with recurrent nonsynonymous mutations and the curated CGC gene list from COSMIC (29). We found recurrent mutations in known cancer genes *TRRAP*, *FAT1, and NCOA1*. We found 3 missense mutations in *TRRAP* (p.Pro252Ser, p.Pro1364Leu, p.Glu2479Lys) across two patients. *TRRAP* is involved a cell-cycle regulation and oncogenesis and has been found to be recurrently mutated in melanoma (45, 46). Notably, previous work showed that skin fibroblasts of individuals with *TRRAP* mutations have significant changes in expression of genes associated with neuronal function and ion transport (45). *FAT1*, a regulator of cell-cell adhesion and extracellular matrix architectural integrity that is a recognized driver of cSCC (47), had one nonsense p.Gln2076Ter mutation and one splice site mutation across two patients. We also found two missense mutations in *NCOA1* (p.Pro1102Ser, p.Ala498Thr) across two PN patients.

We further investigated the mutation rate in known oncogenic pathways as reported by The Cancer Genome Atlas (48). We found the Notch signaling pathway to be most commonly mutated in our PN cohort, with variants detected in 9 out of 17 patients (52.9%), including mutations in *NOTCH1, NOTCH4, CNTN6, FBXW7, JAG2,* and *SPEN*. Notch signaling is followed by the RTK-RAS and Wnt pathways, which were altered in 5 (29.4%) and 4 (23.5%) PN patients, respectively. A list of high-confidence somatic SNV and indel calls made in this study is included in Table S2.

#### Gene set enrichment analysis

Gene set enrichment analysis offers a relatively unbiased view into the somatic landscape at the pathway level and can highlight patterns that are too subtle to identify at the gene level (49). As a proxy for the somatic selection, we first conducted an enrichment analysis of high-VAF (>0.3) nonsynonymous somatic mutations. Different patterns by race were observed (Figure 2). Notably, high-VAF mutations in African American PN patients were associated with epithelial-to-mesenchymal transition (*NOTCH4* and *TASOR*; FDR-adjusted *P* < 0.05).

We then performed enrichment analysis of all genes that were found to have recurrent nonsynonymous somatic mutations in PN lesions. We included pathways from 3 term databases: Gene Ontology (GO) Biological Process, GO Cellular Component, and NCI-Nature Pathways. After correction for multiple hypothesis testing, the most significantly mutated networks were related to Notch-mediated signaling, neuronal migration, and polymeric cytoskeletal fiber (Figure2, B-D).

Since we identified *NOTCH1* as the most frequently mutated gene in PN lesional skin, and loss-of-function mutations in *NOTCH1* have been established to occur commonly in cSCC (50), we compared our PN somatic calls to a publicly available cSCC variant dataset from a recent metanalysis by Chang et al. (47). We included 83 cSCC samples for this analysis, with a median tumor mutational burden and VAF of 21.4 per megabase and 0.258, respectively. Not surprisingly, cSCC samples had a significantly higher mutational burden and VAF than our PN samples (*P* < 0.001, *Wilcoxon*) (Figure 3A and B)*. NOTCH1* was mutated in 55.4% of cSCC samples, second only to *TTN*. Overall, there were 6 genes that mutated in 25% or more of the cSCC samples and also in two or more samples of our PN cohort: *NOTCH1*, *FAT1*, *TTN*, *FLG*, and *RELN* (Figure 3C-E). Out of those genes, *NOTCH1* and *FAT1*, are the ones previously identified as likely cSCC drivers (47) (Figure 3C-E). In addition, *TASOR2,* a gene found to be differentially regulated in several cancers but whose function remains understudied (51), was the only gene not mutated in any of the cSCC samples while having somatic mutations in two of our PN patients.

**Figure 3.**
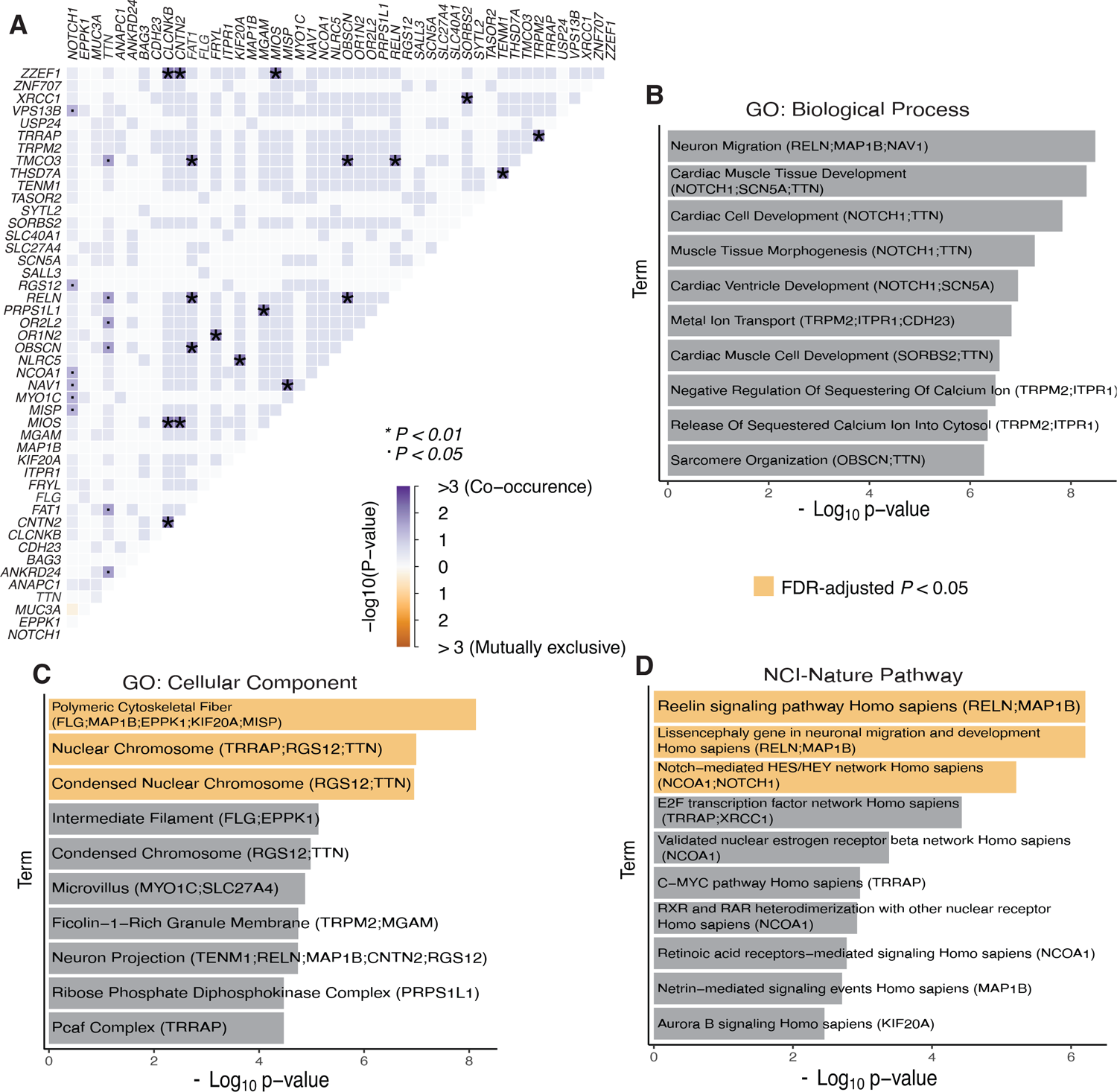
Co-occurrence and enrichment of recurrent somatic mutations in PN lesional skin. (A) Somatic mutational correlation matrix of all 46 genes with recurrent nonsynonymous somatic mutations in PN lesional skin. Significant pairs were identified with Fisher’s exact test. (B-D) Gene ontology (GO) term and pathway enrichment results of recurrently-mutated genes across three pathway databases. Terms with FDR-corrected *P* < 0.05 are colored in yellow.

### Somatic copy number variation in PN

In addition to somatic SNVs and indels, somatic copy-number variation (CNV) may contribute to the phenotype of PN phenotype by altering gene dosage. Copy number analysis of exome data identified a median of 66 somatic CNVs per PN patient (range of 48 to 344), including characteristic recurrent deletions in chromosome 1p13 (6/17; 35%), 4p (5/17; 29%), 4q13.2 (4/17; 24%), 5q (3/17; 18%), 7q21 (3/17; 18%), 11q14 (5/17; 29%), 12 (7/17; 41%), and 19q13.42 (4/17; 24%). Since the method we used gives a greater weight to high amplitude events that are less likely to occur by chance, we also identified significant gains in chromosome 7p22 and chromosome 17q25.3 in one sample (*P* < 0.05; see Methods). We noted distinct patterns of recurrent somatic CNV in PN by race: Caucasian patients more commonly had deletions in chromosome 1p13.3 (4/5; 80%) and duplications in chromosome 15 (3/5; 60%), whereas African American patients more commonly had deletions involving chromosome 4p (5/12; 42%), 4q13.2 (4/12; 33%), 7q21 (3/12; 25%), 11q14 (5/12; 42%), and 12 (5/12; 42%) (Figure 4A-C). Overall, recurrent somatic deletions had an overlap with 3173 gene loci and somatic duplications overlapped with 5 gene loci: *FOXK2*, *WDR45B*, *EIF3B, RSPH10B2*, and *CCZ1B*.

**Figure 4.**
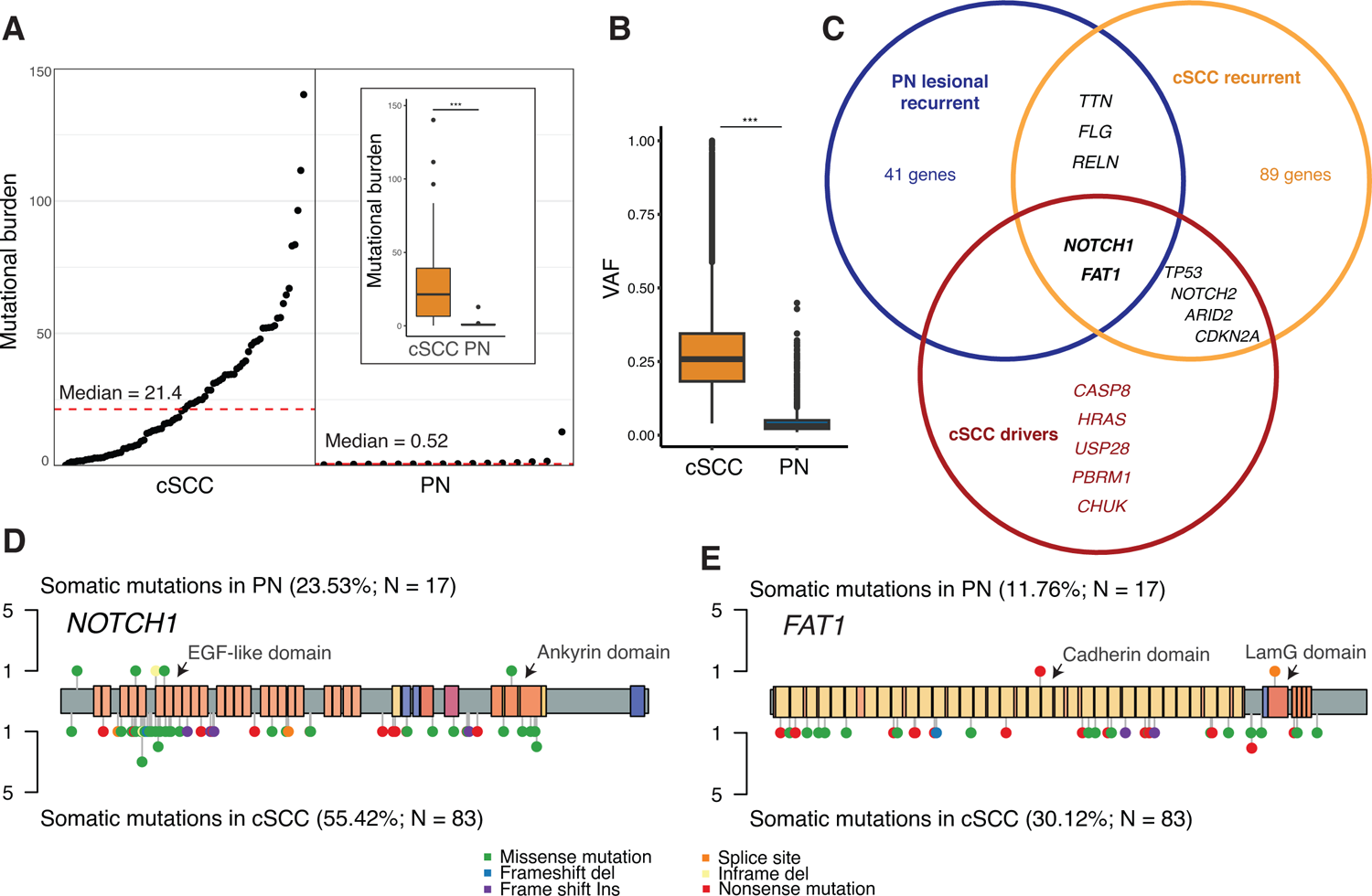
Somatic mutations in PN and cSCC. (A) Rank order plot showing the somatic mutational burden (number of nonsynonymous mutations occurring per megabase of coding regions) across 17 PN patients and 83 cutaneous squamous cell carcinoma (cSCC) patients. Red dashed line indicates the median mutational burden. Inset is a boxplot comparison of somatic mutational burden between the two cohorts. (B) Boxplot showing VAF of all nonsynonymous somatic variants in cSCC and PN patients. (C) Venn diagram depicting the overlap between known cSCC driver genes, genes that were found mutated in 25% or more of the 83 cSCC samples (cSCC recurrent), and genes that were found mutated in 2 or more of our 17 PN samples (PN lesional recurrent). (D, E) Somatic mutations falling within the *NOTCH1* and *FAT1*, respectively. y-axis indicates the number of PN (top) or cSCC (bottom) patients the carrying the somatic mutation. Colored rectangles indicate known functional domains of the protein product. EGF, epidermal growth factor; LamG, laminin G.

Previous studies show that CNV affects gene expression and biological processes (52, 53). To investigate the functional impact of somatic CNV in PN, we overlapped genes affected by recurrent somatic CNV calls in our PN cohort with genes found to be differentially expressed in PN lesional skin using RNA-seq data (10). We identified 264 genes that were significantly downregulated (FDR-adjusted *P* < 0.05; log fold-change < 0) as well as recurrently deleted in PN lesional skin. Enrichment analysis of those 264 genes showed that the deleteriously affected pathways were most significantly related to neural crest cell development, negative regulation of chemotaxis, and negative regulation of axonal extension (Figure 4E). Manual review of the enrichment results revealed that it was primarily driven by a recurrent 7q21 deletion affecting *SEMA3C, SEMA3E, SEMA3A,* and *SEMA3D* (Figure 4D). There were two gene loci that overlapped recurrently duplicated segments and were significantly upregulated in PN lesional skin: *FOXK2* and *EIF3B*.

Since not all of the RNA-seq samples we used overlapped with the WES samples, and to demonstrate that the enrichment in neurotropic pathways is driven by somatic deletions, we repeated the enrichment analysis using expression data only as a negative comparator–there was no significant enrichment in those neuronal pathways without CNV information (Figure S2).

We then performed CNV signature analysis to investigate the common etiology, if any, of such wide-scale somatic events. We identified 3 CNV signatures in our samples based on loss-of-heterozygosity status, total copy number state, and segment length. One CNV signature was highly similar to known signature CN12 from the COSMIC curated database (0.837 cosine similarity), which is believed to be a focal loss-of-heterozygosity signature indicating chromosomal instability in association with a enome doubling event (54). African American and Caucasian patients displayed similar exposure to CN12 (*P* = 0.58, *Fisher’s exact test*) (Figure 4F-G).

### Somatic mutational landscape of PN compared to AD

#### Distinct somatic mutational landscape in PN

Somatic mutational analysis of AD lesional skin revealed a median of 75 lesion-specific mutations per patient (range of 37 to 1161), with the highest frequency of recurrent nonsynonymous somatic mutations in *TTN* (4/9; 44%), *DNHD1* (3/9; 33%), *USP20* (3/9; 33%), and *ANKRD36* (3/9; 33%) (Figure 5 A). Of note, none of the AD samples had somatic mutations in *NOTCH1* (Figure 5D). Somatic mutations in AD lesional skin had a median mutational burden of 0.66 per megabase (range of 0.28 to 92.7) and median VAF of 0.04 (range of 0.010 to 0.405), which were significantly higher than those of PN (*P* < 0.001, *Wilcoxon*) (Figure 5B and E).

**Figure 5.**
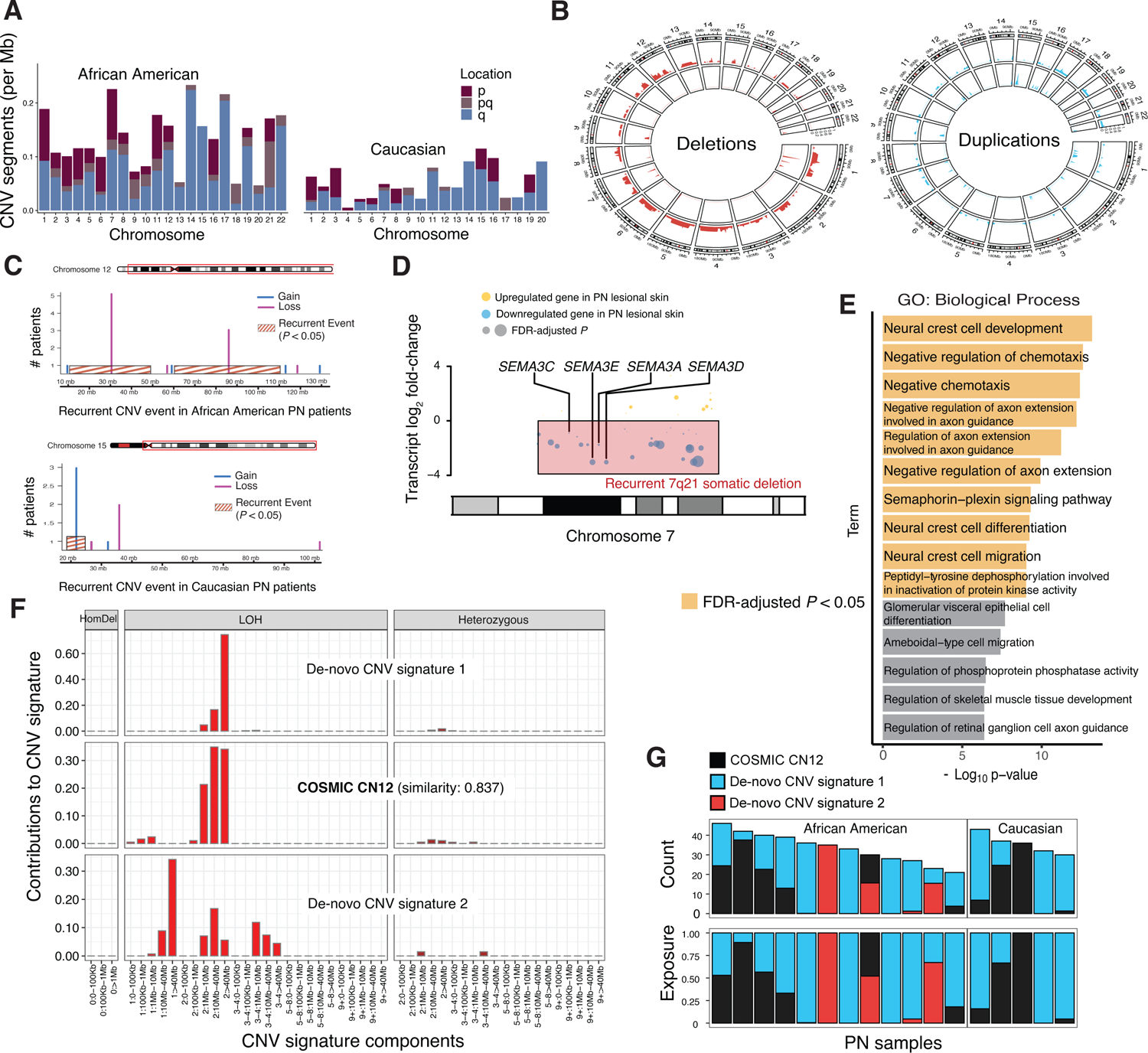
Landscape of somatic CNV in PN. (A) Normalized copy number per sample per chromosome for African American and Caucasian patients. (B) Circular plot illustrating the proportion of samples with deletions (red) and duplications (blue) per genomic region for African American patients (outer ideogram) and Caucasian patients (inner ideogram). (C) Examples of recurrent somatic CNV events only observed in African American patients or Caucasian patients on chromosomes 12 and 15, respectively. (D) RNA-seq data was used to determine differentially down- and up-regulated genes in PN lesional skin. Transcript log2 fold-change is shown on the y-axis with respect to genomic location on chromosome 7 on the x-axis. Overlayed red box shows the location of the recurrent 7q21 somatic deletion. (E) GO term enrichment results of 264 genes that overlapped recurrent deletions while also being differentially downregulated in PN lesional skin. Terms with FDR-corrected *P* < 0.05 are colored in yellow. (F) Decomposition plot showing the relative contributions of CNV signature components to the three somatic CNV signatures detected (HomDel: homozygous deletion, LOH: loss-of-heterozygosity). (G) Distribution of CNV signatures in African American and Caucasian PN patients.

**Figure 6.**
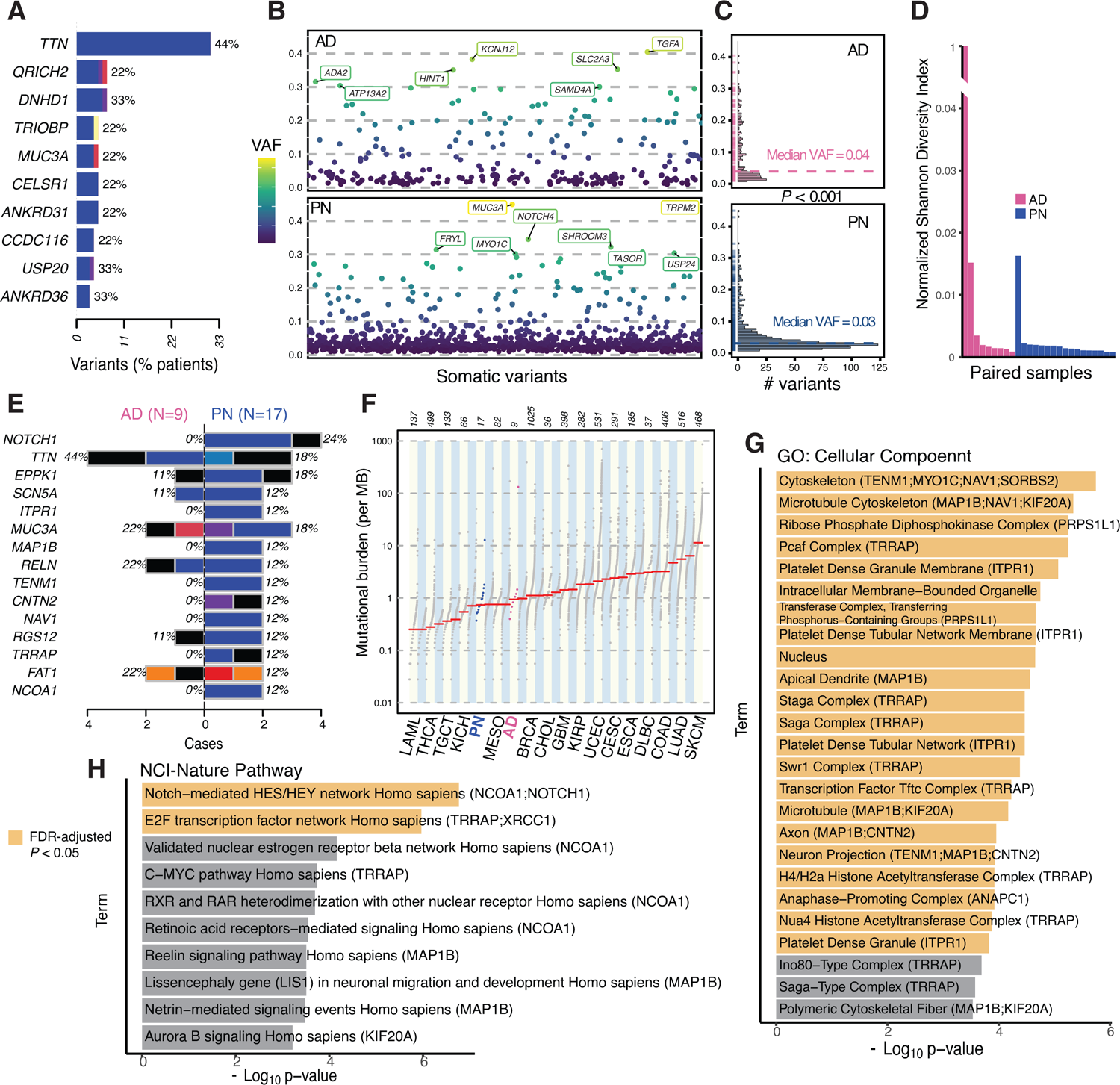
Somatic mutational differences between PN and AD. (A) Ten genes with the most frequent nonsynonymous somatic mutations in our AD cohort. (B) All nonsynonymous somatic mutations in AD and PN are ordered by their genomic locations on the x-axis and the corresponding VAF is shown on the y-axis. Variants with 0.3 or higher VAF are labelled with their gene name. (C) Histograms showing the frequency of nonsynonymous somatic variants on the x-axis at the corresponding VAF on the y-axis, in AD and PN. *P*-value indicates the significant difference in means (*Wilcoxon*). (D) Mutational diversity per sample using a normalized Shannon Diversity Index based on the VAF of somatic SNVs and indels in PN and AD patients. One hypermutated AD sample was not excluded (see Methods). (E) The top mutated genes in our PN cohort are illustrated, with the corresponding relative frequency and percentage of alteration in AD and PN samples. (F) Rank order plot showing the somatic mutational burden per megabase across 17 PN patients, 9 AD patients, and all 33 cancers in The Cancer Genome Atlas cohort. Red lines indicate the median mutational burden. A dictionary for cancer symbols is included in Table S3 (G, H) GO term enrichment results of 21 genes with nonsynonymous somatic mutations in at least two PN samples and no AD samples across two pathway databases. Terms with FDR-corrected *P* < 0.05 are colored in yellow.

To investigate the somatic mutational landscape more characteristic of PN compared to AD, we started with the set of 46 genes that had nonsynonymous mutations in at least two PN patients and filtered those genes that had any nonsynonymous mutations in our AD cohort. This analysis yielded 21 genes for PN. We performed enrichment analysis on the resulting gene list. The most significant pathways were related to the Notch-mediated HES/HEY network, E2F transcription factor network, microtubule cytoskeleton, neuron migration, and others (Figure 5F and G).

We found a nonsense mutation (p.Trp1374Ter) and two missense mutations (p.Glu1017Gly, p.Ser1067Leu) across two AD patients in *DUOX2*. Notably, the same nonsense mutation in *DUOX2* appeared in one PN sample and this is the only recurrent nonsense mutation we observed across both conditions. *DUOX2* is primarily responsible for the release of hydrogen peroxide through NADPH oxidase and variant protein products have been associated with elevated plasma IL17C levels (55, 56).

#### Mutational signatures

Analysis of the mutational processes in PN and AD lesional skin identified the presence of 4 single-nucleotide substitution (SBS) and 2 double-nucleotide substitution signatures (DBS). SBS signatures were most similar to SBS7b (UV exposure; similarity: 0.966), SBS6 (DNA mismatch repair; similarity: 0.806), and SBS5 (unknown etiology, similarity: 0.781), from the COSMIC database (57) (Figure 5A). SBS5 has unknown etiology but it is found to have increased burden in many cancer types and is clock-like in that its exposure correlates with the age of the individual (57). We also detected a de-novo SBS signature in AD and PN characterized by frequent A[T>]G and G[C>T]C substitutions. SBS signatures had similar distribution across PN and AD groups. DBS signatures only correlated with DBS1 (UV exposure; similarity: 0.999), characterized by transcriptional bias with more CC>TT substitutions. We also identified a de-novo DBS signature characterized by frequent TG>CA substitutions. For each detected mutational signature, a multiple linear regression model was built to test if age, sex, PN diagnosis (compared to AD diagnosis), and itch intensity significantly predicted the corresponding signature’s exposure. The regression model of DBS1 was statistically significant (goodness-of-fit adjusted *R*^2^ = 0.66, *P* = 0.004). DBS1 was significantly associated with PN diagnosis (β = 0.779, *P* < 0.001) and itch intensity (β = 0.187, *P* = 0.003).

#### Immunofluorescence analysis

Our somatic analyses identified *NOTCH1* as the most frequently mutated gene in PN lesional skin. As functional validation, we performed immunofluorescent (IF) staining of four lesional and four matching nonlesional PN samples confirmed to have mutations in *NOTCH1*. Representative skin section images are shown in Figure 7A. Intensive staining for the notch intracellular domain (NICD), an indicator of active Notch signaling, was observed in both lesional and nonlesional skin of PN patients. However, NICD showed significantly higher expression in lesional dermis compared to nonlesional dermis from the same PN patients (t(3) = 5.92, *P* = 0.010, *paired Student’s t-test*). There was no significant difference in NICD expression between lesional and nonlesional PN epidermis (t(3) = −0.226, *P* = 0.836) (Figure 7B and C). PN lesional skin had significantly higher co-localization of Notch1 with vimentin, a relatively specific marker for fibroblasts, compared to nonlesional skin of the same patients (t(3) = 4.77, *P* = 0.018) (Fig 7 D & E). While *NOTCH1* also co-localized with KRT10, a keratinocyte marker, there was no significant difference in co-localization between lesional and nonlesional PN skin (t(3) = −2.09), *P* = 0.128) (Figure S2).

**Figure 7.**
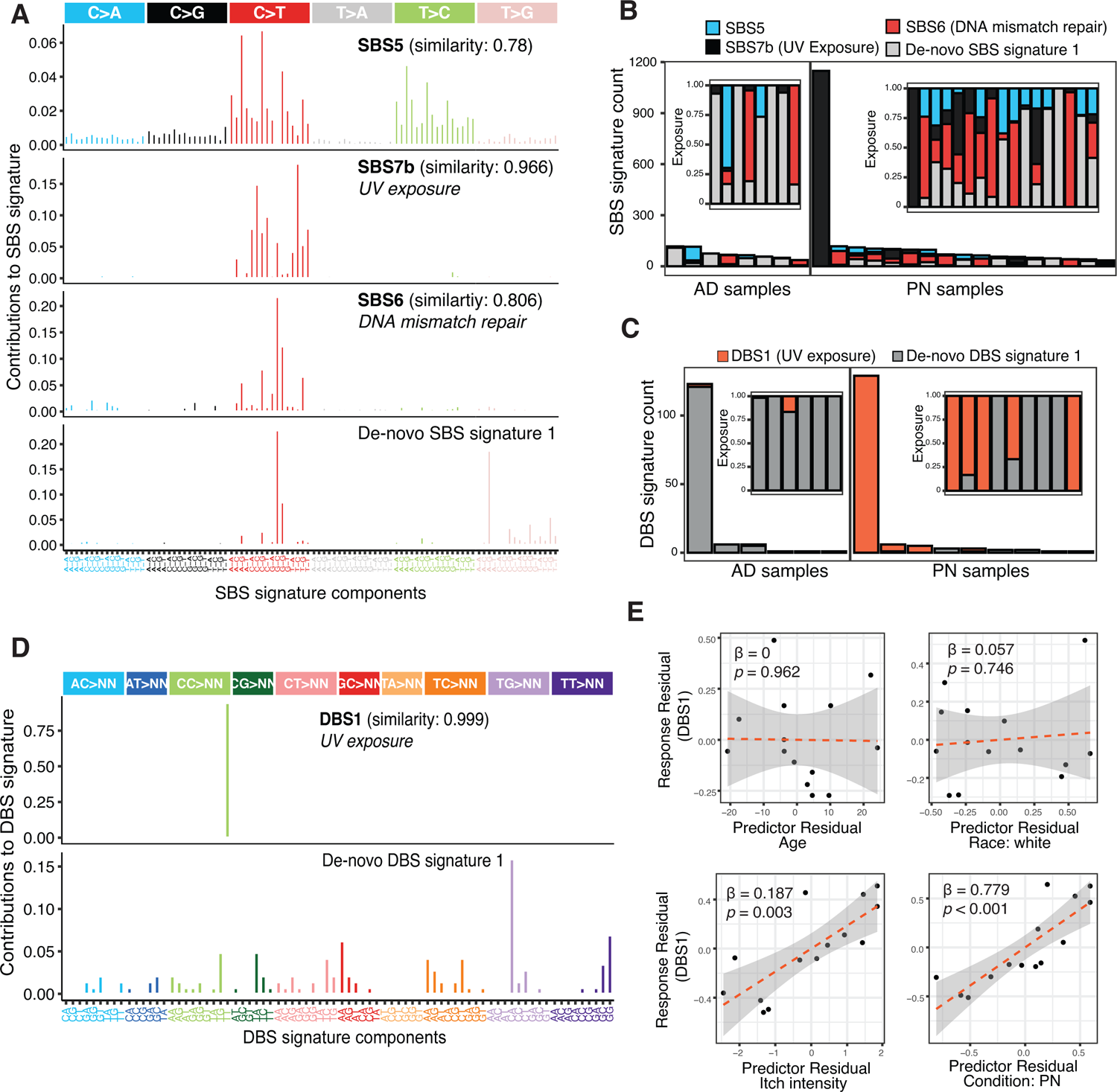
Somatic mutational signatures in PN and AD. (A) Decomposition plot of the 4 somatic SBS signatures detected showing the relative proportion of each transition and transversion subtype. (B) Distribution of SBS signatures in PN and AD samples. The inset figure shows the relative exposures of mutational signature types. Not shown, but included in all analyses, is a hypermutated AD sample with >20,000 instances of SBS5. (C) Boxplot showing the distribution of SBS signatures in PN compared to AD. (D) Decomposition plot of the two somatic DBS signatures detected showing the relative proportion of each base-pair mutation subtype. (E) Distribution of DBS signatures in PN and AD samples. The inset figure shows the relative exposures of DBS signature types. Samples not shown did not display any known DBS signatures (E) Partial regression plots of DBS1 on each of age, race, itch intensity, and condition (PN versus AD), after controlling for the remaining covariates. Overall model adjusted *R*^2^ = 0.66.

#### Multi-center analysis

To test and demonstrate the clinical relevance of our findings for PN patients, we first identified the top 10 non-congenital, nonredundant diseases with the highest evidence for *NOTCH1* involvement, as determined by the gene-disease association (GDA) score from DisGeNET (58). Highest GDAs included aortic valve calcification (0.65), precursor T-cell lymphoblastic leukemia/lymphoma (0.6), and head and neck SCC (0.6). We then leveraged a mult-center cohort through the TriNetX Research Network. 42,397 PN patients without a history of any neoplasms were identified. Controls were identified through 1:1 propensity-score matching based on age, sex, race, ethnicity, smoking status, and history of hypertension (Figure 8A; Table S3). Compared to matched controls, PN patients had a higher relative risk (RR, [95% confidence interval]) of precursor T-cell lymphoblastic leukemia/lymphoma (5.33, [2.88 to 9.88]), head and neck SCC (4.19, [3.22, 5.45]), cervical cancer (3.00, [1.85 to 4.80]), breast cancer (2.91, [2.50, 3.40]), bladder cancer (2.83, [1.924, 4.158]), connective tissue disease (2.48, [2.15, 2.87]), aortic valve calcification (1.96, [1.78, 2.18]), and aortic aneurysm (1.93, [1.680, 2.219]) (Figure 8B). There was no significant increased risk for adenoid cystic carcinoma or glioblastoma in PN patients.

**Figure 8.**
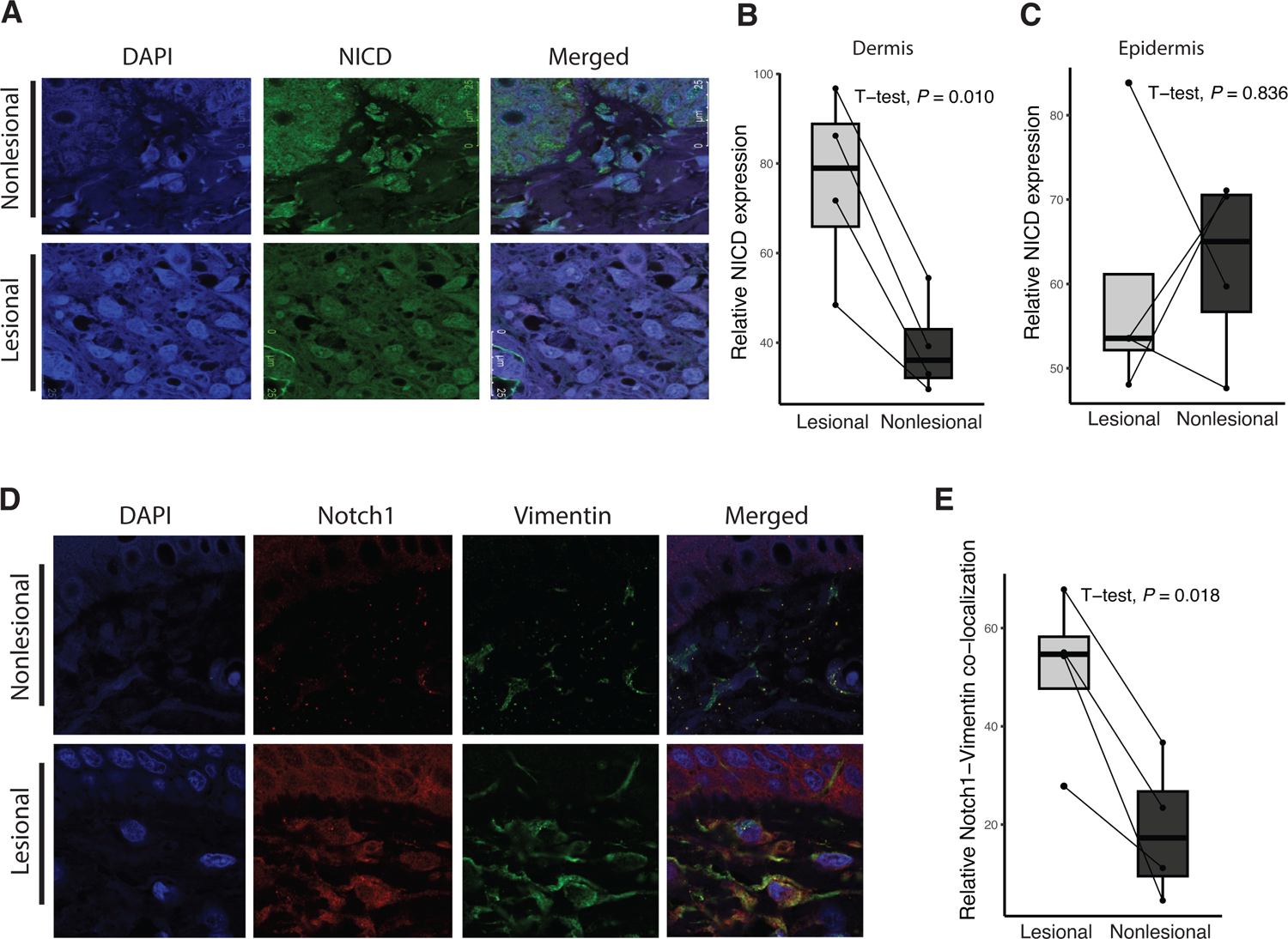
Notch signaling is activated in lesional skin of PN patients. (A) IF staining of NICD in lesional and nonlesional skin sections of a PN patient with a *NOTCH1* somatic mutation. Representative skin sections are shown at 189-fold magnification of the dermis. NICD (green), DAPI (blue). (B & C) Paired boxplot showing the difference in relative expression of NICD between lesional and nonlesional PN dermis (B) and epidermis (C). (D) IF staining of Notch1 and vimentin in lesional and nonlesional skin sections of a PN patient. Notch1 (red), vimentin (green), DAPI (blue). Representative skin sections are shown at 189-fold magnification of the dermis. (E) Paired boxplot showing the difference in relative co-localization of Notch1 and vimentin between PN lesional and nonlesional skin.

**Figure 9.**
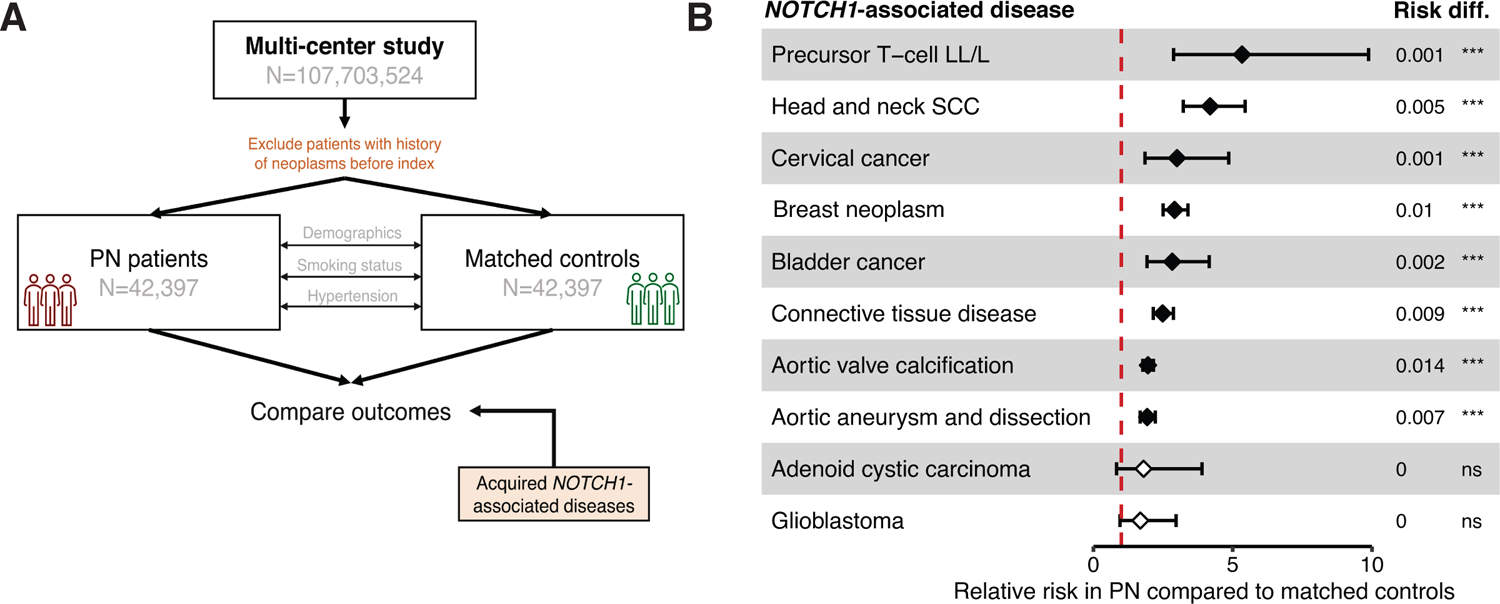
Higher risk of *NOTCH1-*associated diseases in PN patients. (A) A multi-center cohort of PN patients and propensity-score matched controls was obtained using the TriNetX Research Network. The top 10 acquired, non-redundant diseases associated with *NOTCH1* were determined based on available literature support using the DisGeNET database (B) Cumulative relative risk of *NOTCH1-*associated diseases in PN patients compared to matched controls. LL/L, lymphoblastic leukemia/lymphoma; H&N SCC, head and neck squamous cell carcinoma; ***, *P* < 0.0001; ns, no significant difference in risk.

## Discussion

This is the first study to our knowledge to comprehensively investigate somatic mutational events in PN, and we identified recurrent nonsynonymous somatic mutations in PN lesional skin related to the Notch pathway. Increased Notch signaling was observed in PN lesional dermis, along with increased *NOTCH1* expression in fibroblasts from PN lesional skin. PN patients were also found to be at an increased risk of several *NOTCH1-*associated conditions compared to matched controls. Additional findings related to PN biology include downregulated and somatically deleted genes in PN lesional skin associated with neuronal pathways, driven by a 7q21 deletion only seen in African American PN patients. Finally, PN patients had significantly more UV-associated mutational signatures compared to AD patients, after controlling for age and race.

Our findings highlight a role for Notch signaling in PN pathology. The Notch intracellular pathway is highly conserved and plays key roles in specifying cell fates during normal tissue development and homeostasis (59). Abnormal Notch signaling has been implicated in various human diseases and neoplasms (60). Our study identifies *NOTCH1* as the gene with most frequent nonsynonymous somatic mutations in PN. Enrichment analysis revealed that recurrently mutated genes in PN were significantly related to the Notch-mediated HES/HEY network, even after filtering genes mutated in AD controls. *NOTCH1* mutations have been strongly associated with multiple malignancies, including cSCC—a recent metanalysis of 83 cSCCs using multiple cancer gene discovery methods found *NOTCH1* as the top driver gene of cSCC (47). In the present study, *NOTCH1* was the most recurrently mutated gene in PN, and *NOTCH1* mutations significantly co-occurred with *NCOA1* mutations of the same oncogenic pathway, suggesting a proliferative attribute to PN lesional skin. Our comparison between PN and cSCC somatic data also identified *FAT1* as a recurrently mutated gene in our PN samples, another high-confidence driver of cSCC. PN patients are more likely than the general population to have coexisting health conditions, including malignancies (1, 61). In particular, we previously found PN patients at increased risk of developing SCC (25). The present study also found an association between PN and several malignancies using a multi-center cohort, and further found new associations between PN and *NOTCH1*-associated conditions.

In addition to its role in cellular proliferation, Notch signaling also has established roles in tissue fibrosis in several diseases, including renal, hepatic, pulmonary, and myocardial fibrosis (62–65). The profibrotic effect of Notch1 is likely a function of fibroblast proliferation, myofibroblast differentiation, and immune dysregulation through TGF-β signaling (30, 66, 67). Activation of Notch signaling was observed in the lesional skin of patients with systemic sclerosis, and stimulation of healthy dermal fibroblasts with a Notch1 ligand resulted in a phenotype similar to that of systemic sclerosis with increased release of collagen and differentiation of resting fibroblasts into myofibroblasts (68). Interestingly, these findings are aligned with single-cell RNA sequencing studies indicating increased myofibroblasts in PN lesional skin (14).

Given its immunomodulating properties, hyperactive Notch signaling further contributes to fibrosis by inducing an M2 to M1 (proinflammatory) macrophage polarization, leading to myofibroblasts proliferation and recruitment of fibrocytes (69). The transition between M2 and M1 macrophages is also induced by IL-31 (70), which helps connect Notch dysregulation, inflammatory cytokine release, and itch propagation to tissue fibrosis (71, 72). Further, transcriptomic analysis of liver cancer hepatocytes shows a correlation in *NOTCH1* expression and *POSTN*, which encodes periostin (73). Periostin, an extracellular matrix protein released by skin fibroblasts, is found in the dermis of patients with PN at levels that correlate with itch intensity (74). Recent single-cell studies in PN showed significantly increased periostin in lesional skin and supports a fibroblast-neuronal axis in PN regulated by periostin (14). Hyperactivation of a compromised Notch1 receptor may be one possible mechanism by which somatic mutations in *NOTCH1* lead to the clinical cutaneous fibrosis observed in PN lesional skin. IF analysis demonstrated both significantly increased Notch signaling in lesional PN dermis and increased expression of *NOTCH1* in lesional PN skin fibroblasts. Due to its profibrotic and proinflammatory roles, and its relatively high mutation rate and activation in PN lesional skin, our study identifies *NOTCH1* as a driver in PN biology and Notch signaling as a putative target for therapeutics (60).

This study also finds significant somatic mutations in neuronal pathways. Notch1 is known to inhibit neurite outgrowth in neurons, and inhibition of hyperactive Notch signaling can reverse neurogenesis and neurite outgrowth defects (75, 76). We also observed recurrent nonsynonymous mutations in genes related to neuron projection including *MAP1B*, *TENM1*, and *CNTN2*, which were not mutated in AD. Those genes were also found to be significantly downregulated in PN lesional skin compared to nonlesional skin (10), suggesting the mutations lead to altered neuronal structure in PN lesional skin. Further, analysis of somatic CNV in PN reveals that recurrently deleted segments affect genes which are most significantly related to neural crest development and negative regulation of axon extension.

Previous functional studies on lesional and nonlesional PN skin biopsies show altered intraepidermal and dermal nerve fiber density (77). Our mutational profiles suggest primary neuronal gene dysregulation in PN lesional skin. This is supported by association studies showing a correlation between a PN diagnosis and other systemic neuropathies (78).

Following our somatic CNV analysis, enrichment in axonal growth and guidance was largely due to a recurrent 7q21 deletion that overlaps with *SEMA3A* and related genes of the Semaphorin−plexin signaling pathway. This deletion was only observed in African American patients. This is notable in light of the disproportionate burden of PN in skin of color patients, with African Americans having a 3.4 to 4.4 times increased odds of developing PN compared to Whites (26, 61). In addition to CNV differences, we also observed unique high-VAF mutations affecting epithelial-to-mesenchymal transition African American PN patients, suggesting a distinct somatic evolutionary landscape in those patients. This is remarkable given that African Americans with PN often present much differently than Caucasian patients with more fibrotic lesions (Figure 1B and C). Previous transcriptomic studies suggest unique patterns of immune polarization in African American patients with PN (15). Somatic CNV analysis in this study adds further molecular foundation for the high rate of PN in African Americans and suggests a novel disease endotype may exist in these patients.

We observed other mutations that suggest a branch of common etiology between PN and AD. The only recurrent nonsense mutation in PN and AD was in *DUOX2*, which is involved in the release of hydrogen peroxide through NADPH oxidase (55). *DUOX2* is highly sensitive to mutation, and altered protein products have been associated with elevated plasma IL17C levels, which is characteristic of the inflammatory profiles of AD and psoriasis (55). Transcriptional and functional studies show elevated Th17 signatures in PN patients as well, both in the skin and systemically (10, 79). However, the difference in PN is that its immunophenotype is more likely an imbalance between Th17 and Th22, with elevated levels of IL-22 (10). Interestingly, Notch signaling is shown to promote IL-22 secretion and the skewing of naïve CD4^+^T cells toward Th22 cells (80). Further, Notch1 inhibition was shown to effectively alleviate the severity of psoriasis-like skin inflammation by regulating Th17 differentiation and function (81). Hyperactive Notch1 signaling can also destabilize regulatory T cells (82, 83), leaving way for unrestrained Th2-driven inflammation and itch (84). The Notch pathway was found highly mutated and hyperactive in PN lesional in this study, providing support and a genomic context to the Th17/Th22-skewed immunologic signature of PN. This is an area where precision therapeutics will make an impact, as each PN patient’s treatment can be informed by their immunologic or genomic signature.

To investigate the etiology of somatic mutational processes we observed, we performed mutational signature analysis in PN and compared it to that of AD. The relative frequency of DBS1, a highly specific signature for UV exposure with frequent tandem CC>TT mutations, was significantly associated with PN. This was after controlling for age, race, and itch intensity. DBS1 significantly correlated with itch intensity in both conditions. *NOTCH1* is one of the most highly mutated genes in sun-exposed versus non-sun-exposed normal human skin samples (85), and chronic UV-A exposure was shown to expand dermal fibroblasts harboring *NOTCH1* amplifications (86). Subacute skin-barrier damage maybe an early-event in PN patients that increases susceptibility to UV-induced DNA damage, paving the way for accumulated somatic mutations and exacerbating dysregulation in the skin microenvironment. Interestingly, we did not find DBS1 associated with age. Previous work shows a similar pattern in skin fibroblasts where UV-associated DNA damage did not correlate with age, suggesting a proliferative origin (87).

We recognize some limitations in our study. First, we only sequenced samples from 17 PN cases and 10 AD controls, although PN is a relatively rare condition (61). Thus, we may have missed somatic events relevant to PN in a subset of patients. In addition, CNV analysis using WES data is limited in scope and cannot delineate all complex somatic structural variations. Nonetheless, our study represents the first and largest genomic sequencing effort for a rare and understudied disease. Our characterization of the somatic landscape in PN reveals novel insights into its pathology. Aberrant Notch signaling is identified as a likely driver in PN development, likely through profibrotic and immune deregulatory functions, with potential systemic involvement. We also provide support for the neuronal dysregulation pathophysiology of PN through identifying recurrent loss-of-function mutations in *MAP1B*, *TENM1*, and *CNTN2*, as well as recurrent copy number deletions supported by gene expression data. Finally, our mutational signature analysis revealed a strong association between DBS1 and PN, suggesting a potential role for UV-exposure in PN development or maintenance. Our findings represent much needed progress to profile PN on the molecular level.

## Methods

### Sample collection

Patients diagnosed with moderate-to-severe PN, with more than 20 nodules and a Worst-Itch Numeric Rating Scale (WINRS) score (88) of more than 7 out of 10 were recruited from the Johns Hopkins Itch Center. For AD controls, patients diagnosed with moderate-to-severe AD with a validated Investigator Global Assessment (vIGA) score (89) of greater than or equal to 3 and a WINRS score of more than 7 out of 10 were recruited from the Johns Hopkins Itch Center. 6-mm punch biopsies from lesional PN or AD skin and healthy, nonlesional skin within 10 cm of the nodule were collected. Half of each biopsy was formalin-fixed, paraffin-embedded (FFPE) and the other half was stored in RNALater solution (Ambion). All patients signed a consent form approved by the local Institutional Review Board.

### Whole-exome analysis

Library preparation was performed with the SureSelectXT reagent kit before hybridization. SureSelect XT Human All Exon V5 library was used for hybridization. NovaSeq6000 S4 was used for sequencing 150 bp paired-end reads. Illumina’s CASAVA (v1.8.4) was used to convert BCL files to FASTQ files. Initial quality control was performed using FastQC (v0.11.8). Trimgalore (v0.6.7) (90) was used to trim adapters, low-quality base calls, and short reads using default parameters. Following the “Best Practices” workflow suggested by the Broad Institute, BWA-mem (v0.7.17) (91) was used for alignment against the hg38 reference genome, Piccard-tools (v2.9.0) were used to mark duplicate reads, GATK (v3.8.0) IndelRealigner (92) was used to clean indel artifacts, and GATK BaseRecalibrator was used to recalibrate base quality scores and improve downstream variant calling. Samtools (v1.10) (93) and GATK were used to determine coverage at different levels of partitioning and aggregation. One AD sample was excluded due to low coverage (see Results). GATK MuTect2 was used to call somatic. To focus on somatic events relevant to the development of a prurigo nodule, and to further reduce the likelihood of germline calls, MuTect2 paired mode was used with lesional and nonlesional samples as the “tumor” and normal samples, respectively. The default parameters were used. Common germline variants and artifacts were filtered using a panel of normal exomes from the ExAC database; variants present in 2 or more samples within the panel of normals were removed. Somatic variants were then filtered using the following criteria: minimum phred quality of 20, minimum read depth of 20, and minimum variant allele frequency of 0.01. The functional effects of passed somatic SNVs and indels were then predicted using SnpEff (v5.0) (94). The R package Maftools (v2.21.05) (95) was used to summarize and visualize variant calls.

After variant calling, we considered excluding a hypermutated AD sample with 34,228 detected somatic variants after filtration. The rest of the AD samples had a median of 83.5 variants (range of 48 to 578). While this high number of variants might represent significant underlying germline variation or technical artifacts, it may also reflect true hypermutation related to AD. Since this study did not focus on investigating AD pathology, and the AD variants were rather primarily used to curate a gene list that is more likely to be specific to PN somatic mutagenesis, the hypermutated AD sample was not excluded.

### Gene set enrichment analysis

Enrichr (96) was used to perform gene set enrichment analysis using the following term databases: GO Biological Process, GO Cellular Component, and NCI Nature Pathways. Significant GO terms and pathways were calculated using an alpha level of 0.05 after applying the Benjamini–Hochberg correction and output was visualized in R.

### Copy number analysis

Somatic CNV was inferred using CNVkit (v0.9.4) (97), using baited genomic regions for the whole-exome target capture kit S04380110 (i.e. SureSelect Human All Exon V5). Nonlesional samples were combined into a pooled reference for CNV calling, as opposed to an individual-matched analysis, which is the recommendation of the CNVkit authors for reduced CNV noise. Significantly recurrent CNV segments were identified using CNVRanger (98), which implements the statistical approach described by Beroukhim et al. to highlight regions that are aberrant more often than would be expected by chance, with greater weight assigned to high-amplitude events (homozygous deletions or high-level copy-number gains) (99).

*RNA-seq data.* In order to corroborate and contextualize the functional impact of somatic CNV calls, we utilized in-house gene expression data from lesional and non-lesional PN samples. Differential expression pipeline is described previously (100). Briefly, normalization and differential expression of RNA-seq data was carried out using the DESeq2 (101) R package, with adjustment for multiple hypothesis testing using Benjamini-Hochberg. Genes with adjusted-*P* less than 0.05 and log_2_ fold-change change < 0 or > 0 were considered down- and upregulated, respectively. A relatively permissive absolute fold-change cutoff was used to allow enrichment to be driven by somatic CNVs.

### Mutational signature analysis

Mutational signatures for the landscape of single nucleotide substitution (SBS), double nucleotide substitution (DBS), and CNV across exomes were extracted based on the non-negative matrix factorization method previously described and implemented in the Sigminer R package (v2.1.7) (102, 103). CNV signatures were classified as recently described by Steele et al.; in brief, to capture biologically relevant copy number features, a CNV signature encodes the copy number profile of a sample by summing the counts of segments into a 48-dimensional vector based on total copy number, heterozygosity status, and segment size (54). The optimal number of mutational signatures to extract was determined by inspecting the cophenetic correlation coefficient, an indicator of the robustness of consensus matrix clustering, and choosing the minimal number of signatures after which the coefficient starts sharply decreasing (104). Signatures were then compared with the curated set of COSMIC signatures v3.3 (57) using cosine similarity. Signatures with less than 0.7 similarity to any known signatures in COSMIC were considered de-novo.

### Mutational burden

We calculated mutational burden as the number of nonsynonymous mutations occurring per megabase of coding regions.

#### Shannon Diversity Index for somatic mutations

Variant-allele frequency (VAF; *v*) was inferred as the number of reads supporting the variant allele, SNV or indel, divided by the total reads supporting the reference allele and the reads supporting the variant allele. Assuming we sequenced n sites, the Shannon Diversity Index, H, for a sample was then calculated as:

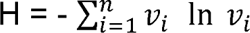

#### Immunofluorescence analysis

We selected four PN patients whom we found to have *NOTCH1* mutations. Lesional and nonlesional FFPE skin samples of 5-µm thickness were deparaffinized and subjected to heat-induced antigen retrieval using Trilogy buffer (Trilogy® 920P x1, Sigma-Aldrich) and treated with DAKO protein blocking reagent (X0909, DAKO, Capinteria, CA, USA). The slides were then incubated with primary antibodies for Notch1 (ab52627, Abcam, 1:150) and human NICD (AF3647, R&D Systems; dilution, 1:20) at 4°C overnight, followed by reaction with conjugated secondary antibodies, DAPI intranuclear stain (62248, Thermofisher Scientific) and mounted with ProLong Glass Antifade Mountant (P36980, Thermofisher Scientific). For quantification, photomicrographs were obtained with Leica SP8 confocal microscope (Leica Microsystems, Deerfield, IL, USA) at 20x objective and x63 objective oil immersion using instrument settings. Background normalized fluorescence intensity of the antibodies in the epidermis and dermis was measured in arbitrary units (AU) using Image J software (NIH, Bethesda, MD, USA).

#### Gene-disease associations

We used DisGeNET (v7.0) (58) to identify diseases associated with *NOTCH1*. At the time of the study, DisGeNET had 1,134,942 gene-disease associations (GDAs) between 21,671 genes and 30,170 diseases. Their methodology ranks diseases based on a GDA score which gives a higher weight to associations reported by several expert-curated databases and with a large number of supporting publications (58). We excluded congenital conditions (e.g., bicuspid aortic valve) and redundant terms (e.g., malignant neoplasm and breast neoplasm: only breast neoplasm was included). The 10 remaining diseases with the highest GDA were then selected as *a priori* primary outcomes in our PN cohort study.

#### Cohort study

The TriNetX Research Network is an international, federated clinical database which contains approximately 107 million patient records at the time of this study. We first identified patients with a diagnosis of PN without any previous history of neoplasms before PN diagnosis. We utilized the International Classification of Diseases 10^th^ Revision Clinical Modification (ICD-10-CM) code L28.1, which is given mostly by dermatologists and has been validated (105). Controls were identified through 1:1 propensity-score matching based on age, sex, race, ethnicity, smoking status, and history of hypertension. Primary outcomes were determined through corresponding ICD-10-CM codes and association was determined through cumulative relative risk estimated using the TriNetX analytics web platform. All TriNetX analyses were completed on 02/10/2023.

#### Statistics

Statistical tests and visualizations were performed using R (version 4.2.0). Adjustment for false-discovery rate (FDR) due to multiple hypothesis testing was conducted using the Benjamini–Hochberg method. An alpha level of 0.05 was used to denote significance.

#### Study approval

This study was approved by the Johns Hopkins Institutional Review Board (IRB00231694).

## Data Availability

All data produced in the present study are available upon reasonable request to the authors.

## Acknowledgements

S.G.K. is supported by the National Institute of Arthritis and Musculoskeletal and Skin Diseases of the National Institutes of Health under Award Number K23AR077073 and a grant from the Skin of Color Society. The content is solely the responsibility of the authors and does not necessarily represent the official views of the National Institutes of Health. We thank the Sidney Kimmel Comprehensive Cancer Center Experimental and Computational Genomics Core, receiving support through NCI grant P30CA006973, for assistance with the WES next generation sequencing studies and analysis. We also thank the PN and AD patients who donated tissue samples for this study.

## Author contributions

SGK, SY, AG, YRS, MMK, TP, and AR designed the study. VP, JD, HC, AK, and SVR collected the data. AR, AG, MDS, and SW analyzed the data. HC, AK, and OO performed staining. AR and SGK wrote the first version of the manuscript. All authors reviewed and edited the manuscript.

## Notes

### Competing Interest Statement

S.G.K. is an advisory board member/consultant for Abbvie, Aslan Pharmaceuticals, Arcutis Biotherapeutics, Celldex Therapeutics, Galderma, Genzada Pharmaceuticals, Incyte Corporation, Johnson & Johnson, Novartis Pharmaceuticals Corporation, Pfizer, Regeneron Pharmaceuticals, and Sanofi and has served as an investigator for Galderma, Pfizer, Incyte, and Sanofi. S.Y. receives research funding to his institution from Bristol-Myers Squibb and Celgene, Janssen, and Cepheid for unrelated work and has served as a consultant for Cepheid. He owns founder's equity in Brahm Astra Therapeutics and Digital Harmonic. All other authors declare they have no competing interests.

### Author Declarations

This study was approved by the Johns Hopkins Institutional Review Board (IRB00231694).

