## Supplemental PDF for "Somatic mutations reveal hyperactive Notch signaling and racial disparities in prurigo nodularis"

**This PDF includes:**

Figures S1-S3
Tables S1, S3, and S4

**Other supplementary material for this manuscript:**

Table S2

**Figure S1**. **Hypermutated PN sample.** (Related to Figure 1)

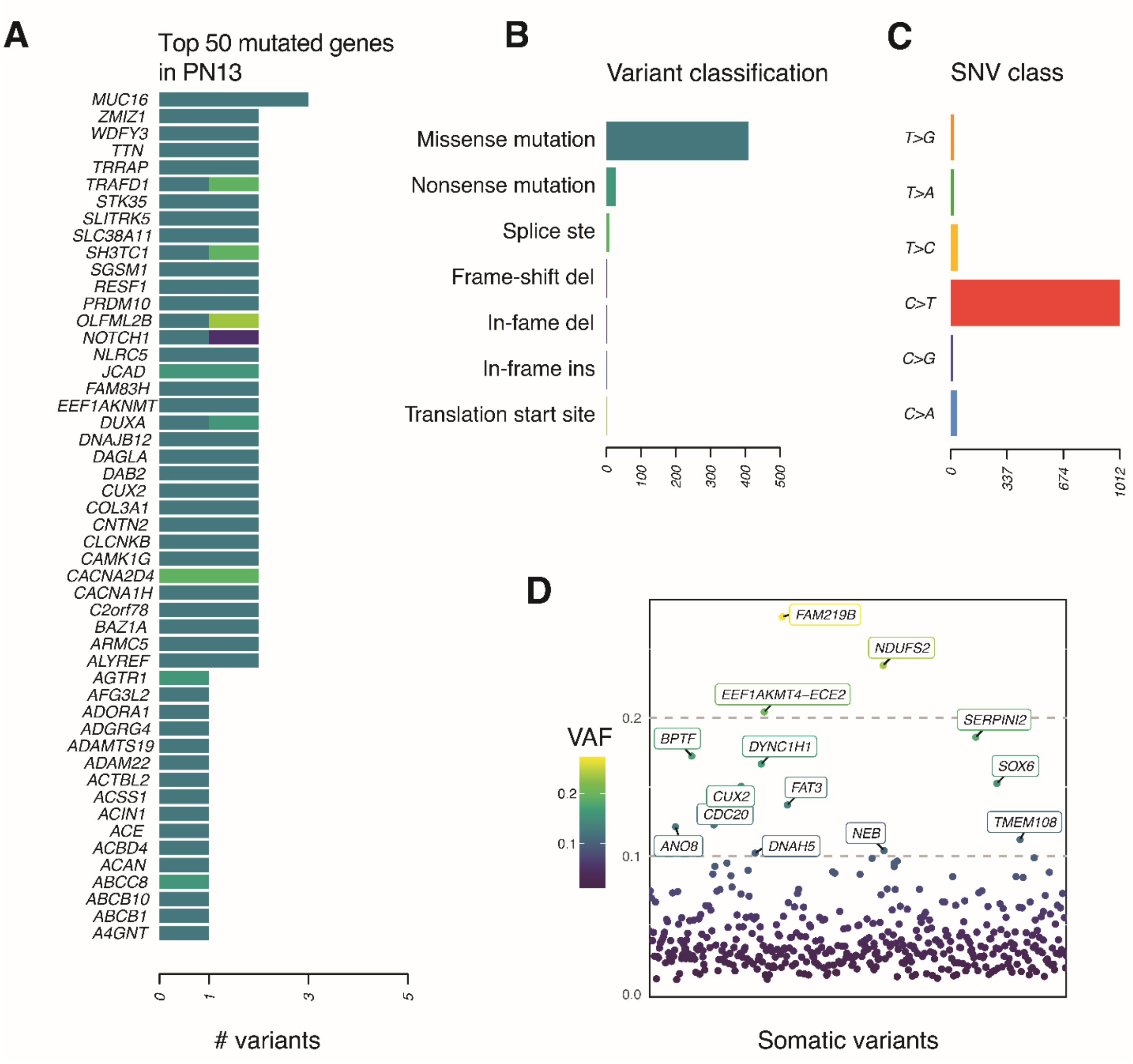


(A) List of the top 50 genes with highest nonsynonymous somatic mutations in hypermutated sample PN13. (B) Frequency of somatic mutations, classified by impact on protein function. (C) Frequency of somatic mutations, classified by single-base substitution type. (D) All nonsynonymous somatic mutations in PN13 are ordered by their genomic locations on the x-axis and the corresponding VAF is shown on the y-axis. Variants with 0.1 or higher VAF are labelled with their gene name.

**Figure S2**. **Gene set enrichment analyses.** (Related to Figures 5 and 6)

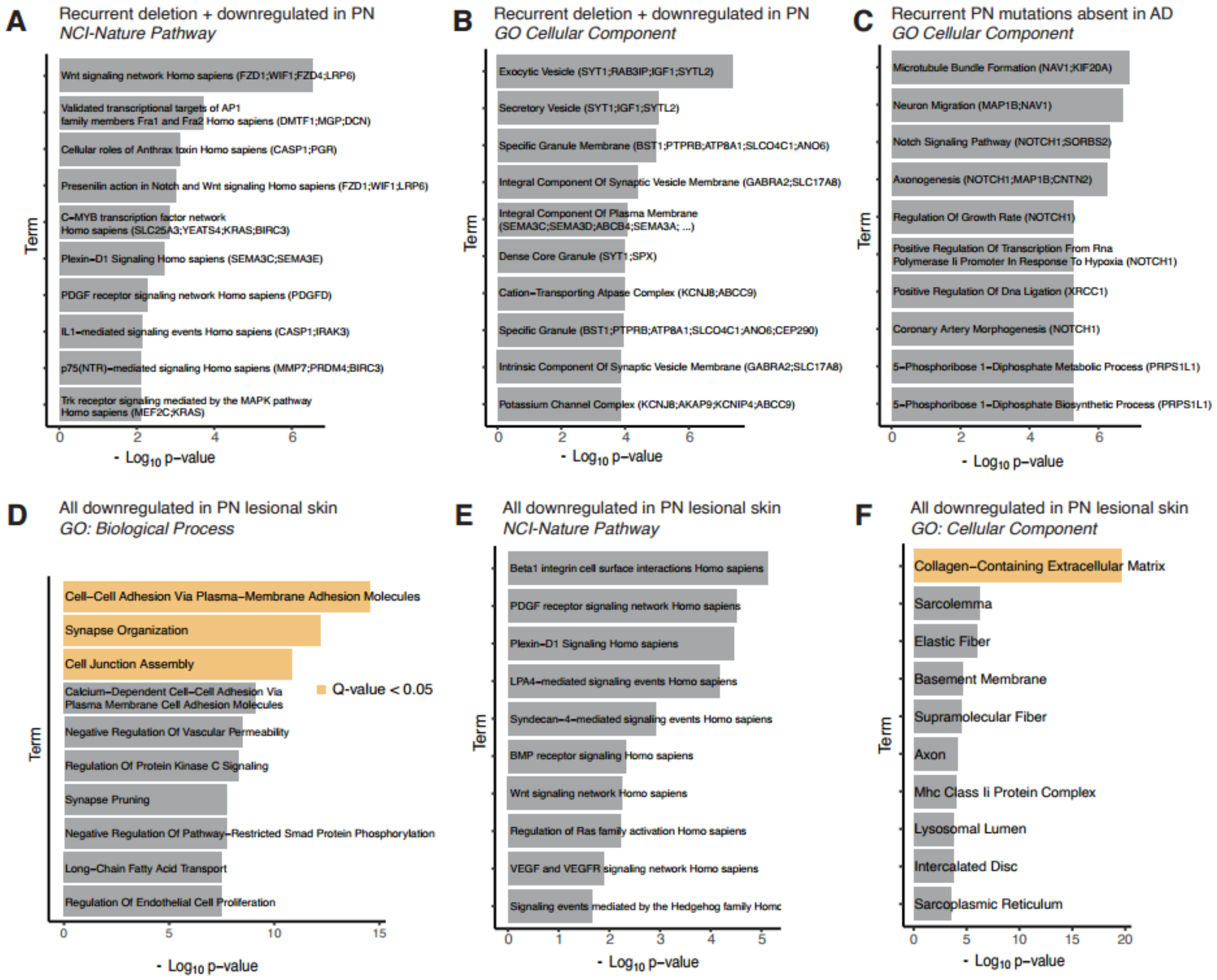


Supplemental enrichment analyses using term databases (NCI-Nature Pathway, GO: Biological Process, or GO: Cellular Component) not included in the corresponding main figures. (A-B) Term enrichment analysis of all recurrently somatically deleted and significantly downregulated genes in PN lesional skin. (C) Term enrichment analysis of 21 genes with recurrent somatic mutations in PN but none in AD. (D-F) We repeated the GO term enrichment analysis (Figure 4E) using all 3,981 genes found downregulated in PN lesional skin (without overlap with recurrent CNV information). Terms with FDR-corrected P < 0.05 are colored in yellow.

**Figure S3. Notch signaling in PN lesional skin.** (Related to Figure 8)

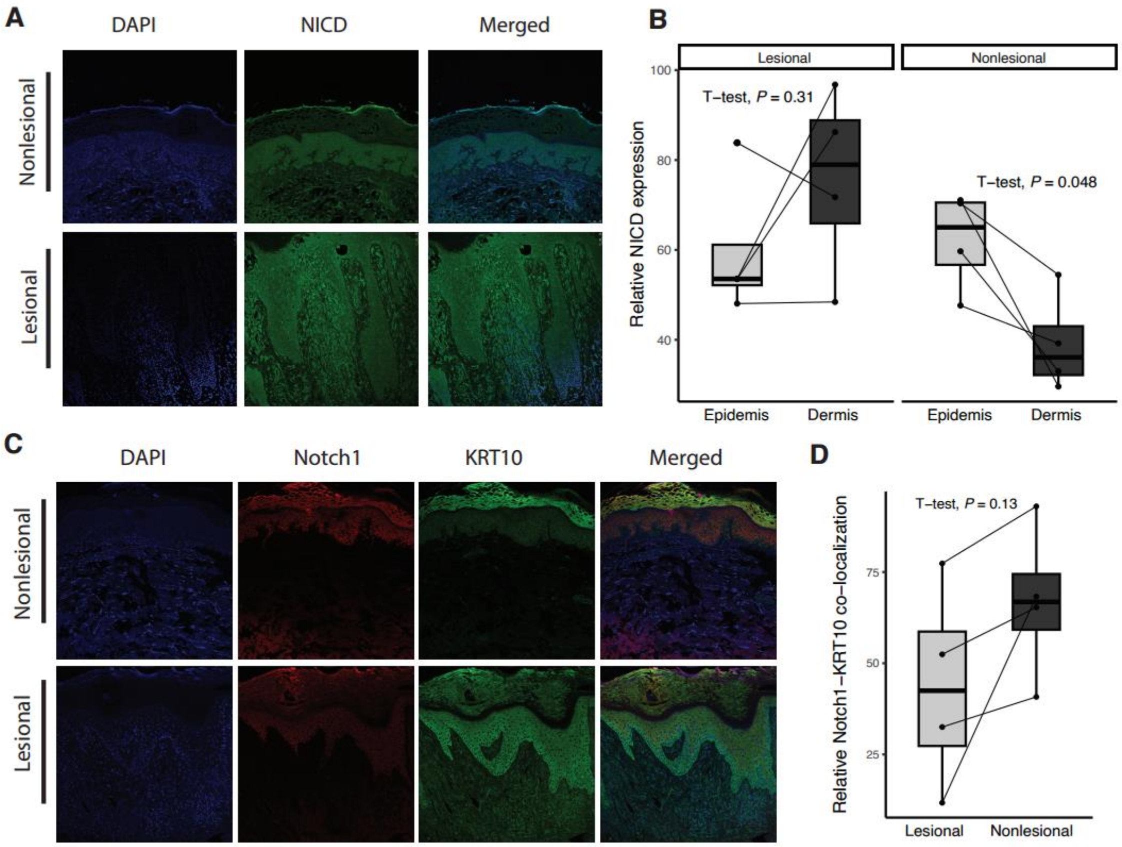


(A) IF staining of NICD in lesional and nonlesional skin sections of a PN patient with a NOTCH1 somatic mutation. Representative skin sections are shown at 40-fold magnification. NICD (green), DAPI (blue). (B) Paired boxplot showing the difference in relative expression of NICD between PN epidermis and dermis in lesional and nonlesional skin. (D) IF staining of Notch1 and KRT10 in lesional and nonlesional skin sections of a PN patient. Notch1 (red), KRT10 (green), DAPI (blue). Representative skin sections are shown at 40-fold magnification. (E) Paired boxplot showing the difference in relative co-localization of Notch1 and KRT10 between PN lesional and nonlesional skin.

**Table S1**. WES and alignment statistics of lesional and nonlesional samples from PN and AD patients.

| Sample name | Total read pairs | Passed read pairs | Aligned read pairs | Average coverage | % base >20X |
|---|---|---|---|---|---|
| AD01_Les | 46,160,417 | 46,142,137 | 46,077,328 | 183.67 | 96.4 |
| AD01_NonLes | 45,678,230 | 45,662,169 | 45,605,145 | 182.82 | 96.4 |
| AD02_Les | 58,683,142 | 58,658,774 | 58,589,249 | 219.94 | 97.1 |
| AD02_NonLes | 47,341,585 | 47,321,736 | 47,199,560 | 185.11 | 96.8 |
| AD03_Les | 41,011,419 | 40,998,518 | 40,945,801 | 165.46 | 95.7 |
| AD03_NonLes | 49,878,240 | 49,860,942 | 49,778,484 | 192.24 | 96.9 |
| AD04_Les | 50,742,258 | 50,709,341 | 50,635,699 | 204.51 | 53.7 |
| AD04_NonLes | 45,693,665 | 45,675,292 | 45,593,139 | 179.16 | 96.1 |
| AD05_Les | 41,445,120 | 41,429,614 | 41,370,519 | 163.26 | 96.2 |
| AD05_NonLes | 49,800,687 | 49,783,616 | 49,686,145 | 192 | 95.5 |
| AD06_Les | 41,386,664 | 41,369,475 | 41,299,752 | 163.8 | 95.4 |
| AD06_NonLes | 65,787,405 | 65,761,670 | 65,668,988 | 241.97 | 97.3 |
| AD07_Les* | 46,401,360 | 46,384,217 | 46,327,118 | 183.71 | 91.1 |
| AD07_NonLes* | 72,787,080 | 72,701,107 | 72,546,883 | 53.18 | 38 |
| AD08_Les | 45,164,463 | 45,151,660 | 45,092,200 | 174.15 | 96.5 |
| AD08_NonLes | 45,390,243 | 45,376,554 | 45,290,384 | 178.13 | 95.9 |
| AD09_Les | 45,260,181 | 45,245,888 | 45,178,713 | 179.05 | 96.2 |
| AD09_NonLes | 42,098,717 | 42,081,084 | 42,006,100 | 164.85 | 96.1 |
| AD10_Les | 40,730,421 | 40,718,688 | 40,111,319 | 156.56 | 94.7 |
| AD10_NonLes | 46,809,990 | 46,796,804 | 46,112,521 | 178.93 | 96.5 |
| PN01_Les | 43,381,478 | 43,366,279 | 43,312,068 | 166.27 | 95.9 |
| PN01_NonLes | 46,282,752 | 46,267,595 | 46,204,071 | 184.69 | 96.3 |
| PN02_Les | 54,976,043 | 54,956,221 | 54,899,077 | 203.57 | 97 |
| PN02_NonLes | 43,732,978 | 43,716,597 | 43,676,125 | 170.4 | 96.1 |
| PN03_Les | 56,006,884 | 55,984,451 | 55,838,863 | 212.06 | 96.9 |
| PN03_NonLes | 52,560,601 | 52,543,300 | 52,429,199 | 206.13 | 96.7 |
| PN04_Les | 44,112,770 | 44,098,137 | 44,028,956 | 165.99 | 96.4 |
| PN04_NonLes | 47,608,236 | 47,590,535 | 47,498,737 | 188.24 | 96.4 |
| PN05_Les | 56,556,972 | 56,534,406 | 56,460,825 | 214.81 | 97.1 |
| PN05_NonLes | 61,487,706 | 61,461,160 | 61,303,243 | 233.73 | 97.5 |
| PN06_Les | 55,350,460 | 55,327,426 | 55,223,154 | 207.25 | 97.2 |
| PN06_NonLes | 56,776,503 | 56,755,447 | 56,685,661 | 221.79 | 97.4 |
| PN07_Les | 56,030,559 | 56,012,202 | 55,884,880 | 210.31 | 96.9 |
| PN07_NonLes | 53,717,595 | 53,695,884 | 53,572,557 | 208.09 | 97 |
| PN08_Les | 41,541,426 | 41,526,931 | 41,479,114 | 161.69 | 87 |
| PN08_NonLes | 48,556,258 | 48,539,888 | 48,468,857 | 187.16 | 93.8 |
| PN09_Les | 44,216,679 | 44,201,428 | 44,137,089 | 166.75 | 96 |
| PN09_NonLes | 49,515,506 | 49,492,487 | 49,406,572 | 203.44 | 70.7 |
| PN10_Les | 48,842,653 | 48,825,855 | 48,769,389 | 185.38 | 95.4 |
| PN10_NonLes | 45,896,235 | 45,862,121 | 45,740,188 | 171.24 | 97.6 |
| PN11_Les | 43,736,958 | 43,718,000 | 43,658,231 | 166.16 | 87.8 |

| | | | | | |
|---|---|---|---|---|---|
| PN11_NonLes | 64,810,969 | 64,770,853 | 64,674,866 | 236.42 | 96.2 |
| PN12_Les | 46,788,922 | 46,762,189 | 46,695,344 | 182.81 | 64.4 |
| PN12_NonLes | 47,106,451 | 47,085,153 | 46,951,000 | 183.59 | 96 |
| PN13_Les | 61,319,095 | 61,292,730 | 61,178,061 | 231.72 | 97.2 |
| PN13_NonLes | 70,449,117 | 70,423,391 | 70,251,859 | 268.64 | 97.7 |
| PN14_Les | 47,644,189 | 47,624,440 | 47,572,723 | 177.18 | 96.3 |
| PN14_NonLes | 50,869,288 | 50,850,639 | 50,762,462 | 201.05 | 96.6 |
| PN15_Les | 45,439,842 | 45,423,436 | 45,362,594 | 175.81 | 96.2 |
| PN15_NonLes | 56,946,952 | 56,924,773 | 56,798,890 | 219.91 | 97.1 |
| PN16_Les | 45,850,373 | 45,834,554 | 45,795,353 | 178.79 | 95.9 |
| PN16_NonLes | 47,672,931 | 47,656,841 | 47,529,912 | 186.9 | 96.7 |
| PN17_Les | 42,740,629 | 42,726,532 | 42,663,696 | 166.86 | 96.1 |
| PN17_NonLes | 43,243,709 | 43,224,747 | 43,165,870 | 169.62 | 96.2 |

*Patient excluded due to low percent of high coverage basepairs in the nonlesional sample.

**Table S2.** All somatic variants identified in lesional PN or AD samples compared to matched nonlesional samples.

Table_S2.xlsx

**Table S3. Dictionary of TCGA cancer symbols.** (Related to Figure 6)

| Symbol | Cancer name |
|---|---|
| ACC | Adrenocortical carcinoma |
| BLCA | Bladder urothelial carcinoma |
| BRCA | Breast invasive carcinoma |
| CESC | Cervical squamous cell carcinoma and endocervical adenocarcinoma |
| CHOL | Cholangiocarcinoma |
| COAD | Colon adenocarcinoma |
| DLBC | Lymphoid neoplasm diffuse large b-cell lymphoma |
| ESCA | Esophageal carcinoma |
| GBM | Glioblastoma multiforme |
| HNSC | Head and neck squamous cell carcinoma |
| KICH | Kidney chromophobe |
| KIRC | Kidney renal clear cell carcinoma |
| KIRP | Kidney renal papillary cell carcinoma |
| LAML | Acute myeloid leukemia |
| LGG | Brain lower grade glioma |
| LIHC | Liver hepatocellular carcinoma |
| LUAD | Lung adenocarcinoma |
| LUSC | Lung squamous cell carcinoma |
| MESO | Mesothelioma |
| OV | Ovarian serous cystadenocarcinoma |
| PAAD | Pancreatic adenocarcinoma |
| PCPG | Pheochromocytoma and paraganglioma |
| PRAD | Prostate adenocarcinoma |
| READ | Rectum adenocarcinoma |
| SARC | Sarcoma |
| SKCM | Skin cutaneous melanoma |
| STAD | Stomach adenocarcinoma |
| TGCT | Testicular germ cell tumors |
| THCA | Thyroid carcinoma |
| THYM | Thymoma |
| UCEC | Uterine corpus endometrial carcinoma |
| UCS | Uterine carcinosarcoma |
| UVM | Uveal melanoma |

**Table S4.** Basic characteristics of the PN and matched control patients in the TriNetX Research Network cohort study. (Related to Figure 9). Patients with prior neoplasms were excluded.

| Cohort | Characteristic | Mean ± SD | Patients | % of cohort | P-value |
|---|---|---|---|---|---|
| PN | Age at Index | 50.2 +/- 18.2 | 42,397 | 100% | 0.985 |
| Controls | | 50.2 +/- 18.2 | 42,397 | 100% | |
| PN | White | | 22,986 | 54.20% | 0.995 |
| Controls | | | 22,985 | 54.20% | |
| PN | American Indian or Alaska Native | | 192 | 0.50% | 0.878 |
| Controls | | | 189 | 0.40% | |
| PN | Female | | 25,665 | 60.50% | 0.961 |
| Controls | | | 25,672 | 60.60% | |
| PN | Native Hawaiian or Other Pacific Islander | | 60 | 0.10% | 0.928 |
| Controls | | | 61 | 0.10% | |
| PN | Unknown Ethnicity | | 8,385 | 19.80% | 0.979 |
| Controls | | | 8,382 | 19.80% | |
| PN | Not Hispanic or Latino | | 30,596 | 72.20% | 0.994 |
| Controls | | | 30,597 | 72.20% | |
| PN | Hispanic or Latino | | 3,416 | 8.10% | 0.98 |
| Controls | | | 3,418 | 8.10% | |
| PN | Black or African American | | 9,259 | 21.80% | 0.993 |
| Controls | | | 9,258 | 21.80% | |
| PN | Male | | 16,725 | 39.40% | 0.961 |
| Controls | | | 16,718 | 39.40% | |
| PN | Unknown Race | | 8,247 | 19.50% | 1 |
| Controls | | | 8,247 | 19.50% | |
| PN | Asian | | 1,653 | 3.90% | 0.943 |
| Controls | | | 1,657 | 3.90% | |
| PN | Nicotine dependence, cigarettes | | 942 | 2.20% | 0.981 |
| Controls | | | 941 | 2.20% | |
| PN | Hypertensive diseases | | 11,289 | 26.60% | 0.994 |
| Controls | | | 11,288 | 26.60% | |
